# GEN-KnowRD: Reframing AI for Rare Disease Recognition

**DOI:** 10.64898/2026.03.02.26347469

**Authors:** Chao Yan, Wu-Chen Su, Yi Xin, Monika E. Grabowska, Vern E. Kerchberger, Victor A. Borza, Jinlian Wang, Liwei Wang, Rui Li, Jacob Lynn, Alyson L. Dickson, Cathy Shyr, QiPing Feng, Charles M. Stein, Kai Wang, Peter J. Embi, Bradley A. Malin, Hongfang Liu, Wei-Qi Wei

## Abstract

Rare diseases affect over 300 million people worldwide, yet patients often endure years-long diagnostic delays that limit timely intervention and trial opportunities. Computational rare disease recognition (RDR) remains constrained by knowledge resources that are often incomplete, heterogeneous, and dependent on extensive multi-disciplinary expert curation that cannot scale. Large language models (LLMs) applied directly for end-to-end diagnosis or disease discrimination face similar knowledge bottlenecks while also raising concerns around cost, reproducibility, and data governance. Here, we introduce GEN-KnowRD, a knowledge-layer-first framework that leverages LLMs to generate schema-guided rare disease profiles, systematically assesses their quality, and constructs a computable knowledge base (PheMAP-RD) for local deployment. GEN-KnowRD integrates this knowledge into lightweight inference pipelines for both general-purpose disease screening and specialized early discrimination from longitudinal electronic health records. Across six public benchmarks for general-purpose screen (9,290 patients spanning 798 rare diseases), GEN-KnowRD significantly improves disease ranking compared to a state-of-the-art, HPO-centered diagnostic framework (up to 345.8% improvement in top-1 success), advanced end-to-end LLM reasoning (up to 129.1% improvement), and a variant of GEN-KnowRD instantiated with expert-curated knowledge rather than LLM-generated profiles. In two real-world cohorts for early diagnosis of idiopathic pulmonary fibrosis (511 patients) as a use case, GEN-KnowRD also demonstrates robust discrimination performance gains, supporting effective RDR during the pre-diagnostic window. These findings demonstrate that repositioning LLMs from diagnostic reasoning to the knowledge layer—decoupling knowledge construction from patient-level inference—yields stronger RDR, while providing scalable, continuously updatable, and reusable infrastructure for diagnosis, screening, and clinical research across the rare disease landscape.

## Main

Rare diseases collectively affect an estimated 300 million people worldwide,^1^ yet their clinical complexity and rarity leave most clinicians with limited experience in recognizing them.^2–4^ As a result, patients often endure years-long diagnostic odysseys, missing critical windows for treatment, clinical trial participation, and therapeutic development.^5^ Computational approaches to rare disease recognition (RDR) have sought to close this gap by extracting phenotypic (and, where available, genotypic) features from patient records, mapping them to standardized representations such as Human Phenotype Ontology (HPO) terms, and matching them against human-curated knowledge resources^6–17^ to prioritize candidate diseases for investigation. However, the knowledge resources this paradigm depends on are typically heterogeneous, incomplete, biased toward well-studied conditions, and reliant on extensive manual curation that is expensive to maintain and cannot keep pace with rapidly evolving medical evidence^18–24^, which increasingly constrain the reliability, scalability, and equity of RDR.

Recent advances in large language models (LLMs) have sparked enthusiasm for end-to-end diagnostic reasoning.^25–29^ However, directly applying LLMs as diagnosticians or discriminators does not fundamentally resolve the knowledge infrastructure problem. The knowledge encoded in LLMs’ parameters is implicit and difficult to govern or trace, while retrieval-augmented approaches remain only as reliable as the underlying corpora and the retrieval pipelines,^30–33^ offering no assurance that the evidence most relevant to a given patient in terms of semantics and granularity will be reliably retrieved,^34^ correctly grounded,^35^ or consistently applied.^36^ These limitations are compounded by practical constraints. Deploying frontier LLMs for routine clinical quires is computationally and financially prohibitive at health-system scale, and transmitting sensitive patient information to proprietary models or external services they invoke raises concerns around data privacy and governance.^37^ Together, these factors motivate an alternative strategy, one that harnesses LLM capabilities upstream, while keeping patient-level inference lightweight, local, and reliable.

We hypothesize that the core limitation of computational RDR lies not in the sophistication of downstream inference but upstream, in how disease knowledge is synthesized, structured, and made computable. Rather than using LLMs as per-patient reasoners that reconstruct knowledge context for each patient-level inference, we reposition them to the knowledge layer where they generate reusable disease representations. Under this paradigm, inference can be executed through lightweight, deterministic pipelines grounded in a shared knowledge layer, thereby decoupling knowledge synthesis from patient-level reasoning.

Here, we present GEN-KnowRD (GENerative Knowledge-driven Rare Disease recognition framework; **Fig. 1a**), a framework that transforms LLM use for RDR by moving LLMs from the point of diagnosis or discrimination to the knowledge layer. Using the rare disease report catalog of the National Organization for Rare Disorders (NORD) database^38^, GEN-KnowRD 1) prompts multiple advanced LLMs (Claude Sonnet 4^39^, DeepSeek R1^40^, Gemini 2.5 Pro^41^, and OpenAI o3^42^) to produce schema-guided disease profiles; 2) conducts multi-faceted quality assessment of the resulting disease profiles; and 3) converts them into a computable knowledge base, PheMAP-RD.

**Fig. 1:**
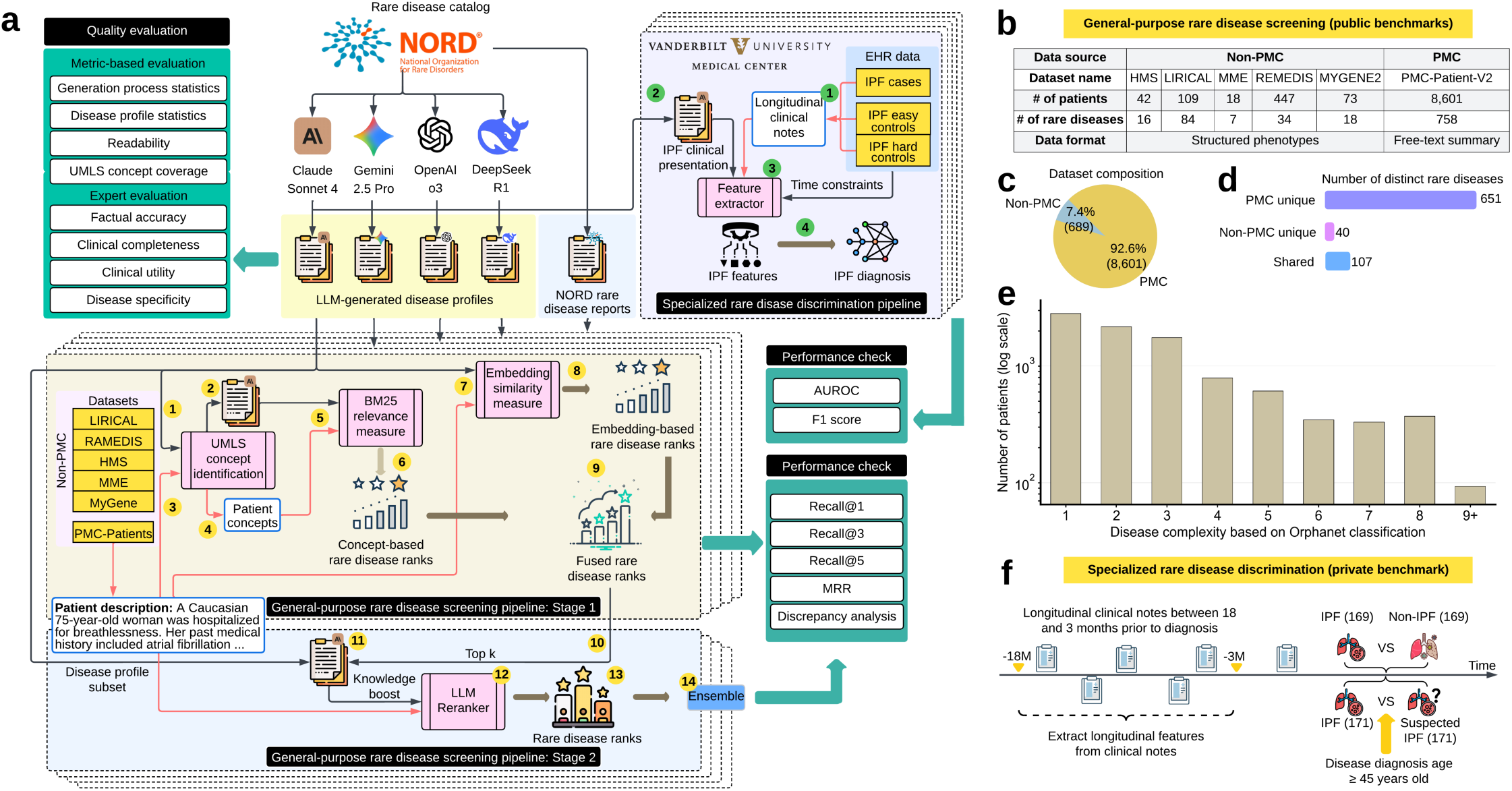
An overview of GEN-KnowRD architecture, evaluation strategy, and datasets used. **a,** For each rare disease in the NORD rare disease report catalog, multiple large language models (LLM) are independently prompted to generate a schema-guided disease profile comprising ten clinically meaningful sections; the quality of LLM-generated rare disease profiles is assessed using a multi-faceted framework that combines objective quantitative metrics with clinical expert review; a general-purpose rare disease screening pipeline then compares both sparse and dense representations of patient clinical presentation description against the corresponding representations of rare diseases to derive an initial disease ranking (Stage 1), followed by a knowledge-boosted reranker that refines the top candidate list (Stage 2); a specialized rare disease discrimination pipeline, illustrated using IPF as a use case, extracts disease-relevant clinical evidence and quantifies symptom presence and severity from longitudinal clinical notes prior to diagnosis, followed by lightweight classification models that distinguish IPF from non-IPF patients and from suspected IPF patients. **b,** A summary of the public benchmarks used to evaluate the performance of the general-purpose rare disease screening pipeline, as well as the utility of disease profiles for rare disease recognition. **c,** Composition of the public benchmarks used for evaluation. **d,** Disease distribution across the public benchmarks after excluding patients whose diseases fall outside of the scope of the NORD report catalog. **e,** Distribution of disease complexity across the public benchmarks, based on the Orphanet disease classification and defined by the number of affected body systems. **f,** A summary of the private benchmark and tasks from Vanderbilt University Medical Center used to evaluate the performance of the specialized rare disease discrimination pipeline, as well as the utility of disease profiles for rare disease recognition. NORD: National Organization for Rare Disorders; LLM: large language model; UMLS: unified medical language system; AUROC: area under the receiver operating characteristic curve; MRR: mean reciprocal rank; IPF: idiopathic pulmonary fibrosis.

Building on this resource, GEN-KnowRD then implements two inferential pipelines that enable rich clinical evidence distillation: one designed for general-purpose disease screening that produces a ranked list of candidate diseases, and another tailored to specialized disease discrimination that classifies whether a patient has a target disease. In our design, these pipelines operate on patient-level clinical evidence derived from existing health records, while PheMAP-RD is applied as a reusable, local knowledge resource to the patient data without requiring patient information to be sent to any LLM. Extensive evaluations across both public (9,290 patients spanning 798 distinct rare diseases) and private (longitudinal cohorts for idiopathic pulmonary fibrosis diagnosis that involve 511 patients from Vanderbilt University Medical Center) benchmarks demonstrate that GEN-KnowRD consistently 1) outperforms a state-of-the-art, two-stage HPO-centered diagnostic framework (PhenoBrain^43^), 2) surpasses pipelines based on human-curated knowledge bases (i.e., NORD rare disease reports^38^), and 3) achieves performance that is better than OpenAI GPT-5, a state-of-the-art LLM baseline performing end-to-end inference. Most notably, GEN-KnowRD based on the knowledge source generated by Claude Sonnet 4 delivers the strongest overall performance and robust gains, surpassing alternative LLM-generated disease profiles across evaluation settings. We have released PheMAP-RD through our public service platform^44^ (Supplementary **Figs. 1,2**) as a reusable resource that can be readily integrated into future diagnostic systems, including retrieval-augmented and agentic LLM pipelines. Rather than competing with end-to-end LLM-or agent-based diagnostic approaches, GEN-KnowRD complements them by providing better-structured knowledge and principled processing modules that can strengthen evidence retrieval, grounding, and downstream decision support.

**Fig. 2:**
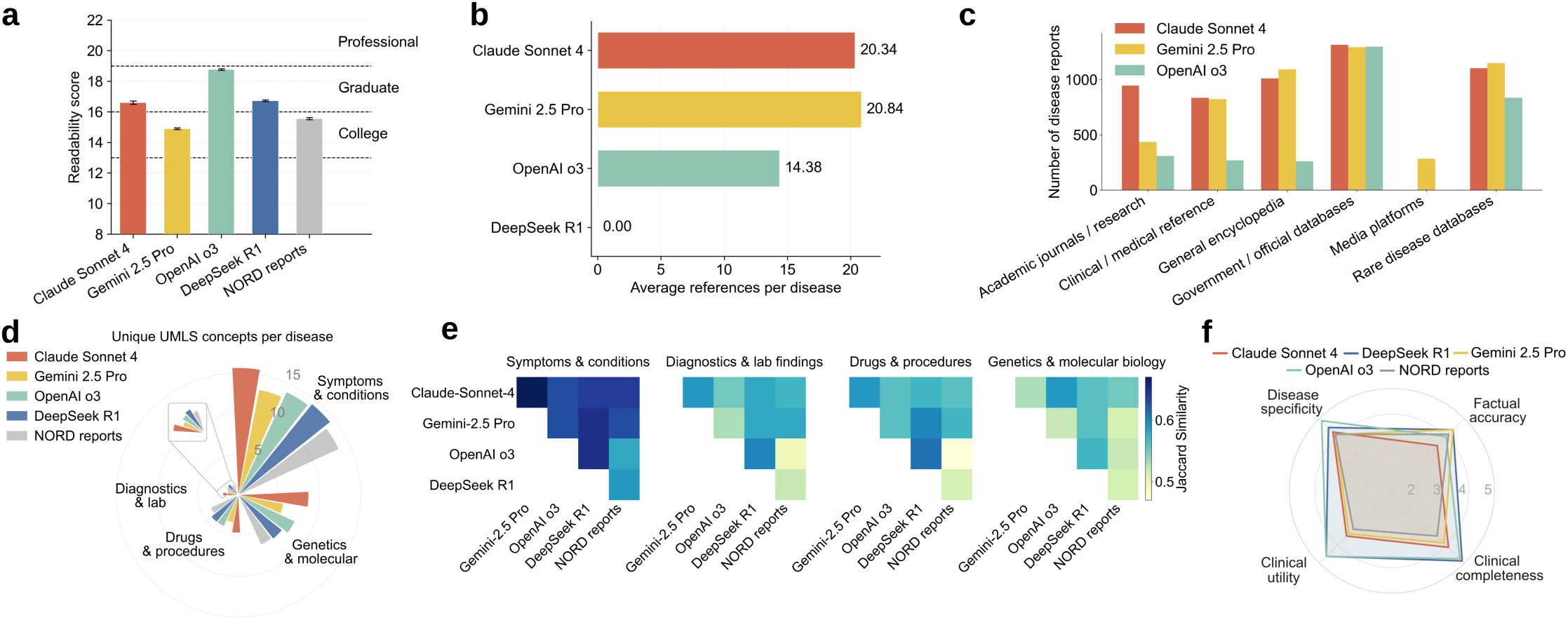
Evaluation of LLM-generated RDK using semantic-based analysis and expert review. **a,** Readability of rare disease profiles measured by the Simple Measure of Gobbledygook (SMOG). **b,** Average number of references generated per disease. **c,** Distribution of reference source types. **d,** Number of unique matched UMLS concepts per disease across semantic groups (each concept is counted once per profile, even if matched multiple times). **e,** Jaccard similarity of UMLS concept sets among models within each semantic group. **f,** Expert evaluation of rare disease profiles for four randomly sampled rare diseases, with scores averaged across two clinical experts blinded to the source of disease profiles.

### An overview of GEN-KnowRD and datasets

GEN-KnowRD is comprised of four components (**Fig. 1a**): 1) LLM-based rare disease profile creation, 2) multi-faceted RDK quality evaluation, 3) general-purpose rare disease screening, and 4) specialized rare disease discrimination. Rather than relying on the knowledge encoded in LLMs or on existing external knowledge for RDR, GEN-KnowRD harnesses LLMs to construct systematic, well-structured, and reusable RDK layer in a machine-actionable form. We scope the rare disease profile creation to the 1,320 diseases in the NORD Rare Disease Database^38^ (which incorporates knowledge from OMIM and Orphanet) for which NORD provides expert-curated, in-depth disease reports covering disease characteristics, diagnosis evaluation, management, and prognosis. This subset serves as a rigorously defined evaluation backbone for three reasons. First, these diseases are supported by structured, clinically vetted descriptions that enable systematic comparison between LLM-generated and expert-curated knowledge under controlled conditions. Second, they correspond to rare diseases with sufficiently annotated patient cohorts across public benchmarks, permitting reliable, label-verified performance evaluation. Third, constraining the candidate space during initial validation allows us to isolate the impact of knowledge-layer quality and inference architecture without confounding effects from diseases lacking standardized descriptions or benchmark cases. Importantly, GEN-KnowRD is modular and not restricted to this subset. The framework can be extended to additional rare or common diseases as structured knowledge sources and labeled cohorts become available. The present scope therefore provides a high-quality, controlled testbed to evaluate knowledge-layer construction and its downstream impact, rather than a structural limitation of the approach.

We selected four representative LLMs, i.e., Claude Sonnet 4^39^, DeepSeek R1^40^, Gemini 2.5 Pro^41^, and OpenAI o3^42^, because they demonstrate state-of-the-art performance across diverse model families and offer strong reasoning and synthesis capabilities well-suited for knowledge layer construction. Each model was then instructed to generate schema-guided multi-section profile for each disease. The profiles cover ten prespecified axes: 1) disease overview, 2) synonyms and abbreviations, 3) subtypes or variants, 4) epidemiology, 5) etiology and pathogenesis, 6) clinical presentation, 7) diagnostic evaluation, 8) management and standard therapy, 9) investigational or emerging therapies, and 10) prognosis. The prompt template used is detailed in Supplementary **Table 1**.

**Table 1:** Summary of LLM-produced rare disease profiles and their generation process.

| Source | Running time (seconds) |  | Input Tokens |  | Output Tokens |  | Reasoning Tokens |  | Reasoning Ratio | Queries per Disease* |  |
| --- | --- | --- | --- | --- | --- | --- | --- | --- | --- | --- | --- |
|  | Mean (STD) | Median (IQR) | Mean (STD) | Median (IQR) | Mean (STD) | Median (IQR) | Mean (STD) | Median (IQR) |  | Mean (STD) | Median (IQR) |
| Claude Sonnet 4 | 144.71 (35.48) | 140.31 (124.97, 160.24) | 323,766 (106,225) | 314,579 (239,060, 401,724) | 5,230 (964) | 5131 (4559, 5811) | NA | NA | NA | 6.10 (1.14) | 6 (5,7) |
| Gemini 2.5 Pro | 87.93 (23.9) | 81.62 (69.98, 102.14) | 1,403 (121) | 1,405 (1,346, 1,472) | 2,645 (554) | 2,607 (2,250, 2,959) | 790 (808) | 430 (275, 988) | 29.9% | 13.36 (7.33) | 11 (9, 17) |
| OpenAI o3 | 88.68 (41.51) | 81.58 (66.20, 104.46) | 157,212 (79,574) | 144,767 (101,700, 195,678) | 3,439 (597) | 3,385 (3,032, 3,801) | 1,288 (444) | 1,216 (960, 1,536) | 37.5% | 9.34 (2.93) | 9 (7, 11) |
| DeepSeek R1 | 141.69 (47.04) | 142.91 (102.19, 171.12) | 749 (3) | 748 (747, 750) | 2,886 (880) | 3,026 (2,031, 3,486) | 769 (590) | 526 (421, 716) | 27.0% | 0 (0) | 0 (0,0) |

NA: not available; STD: standard deviation; IQR: interquartile rage; \*: queries are performed by LLMs to seek for additional information.

Based on the disease profiles generated from each model, we then completed the corresponding RDK by extracting the disease-relevant clinical concepts. Unlike prior research that has relied primarily on HPO terms for RDR tasks^43^, GEN-KnowRD extracts the unified medical language system (UMLS) concepts of selected semantic categories from the collected rare disease profiles, enabling a more informative representation of rare diseases. The rationale for extracting UMLS concepts is twofold. First, HPO is designed to represent phenotypic abnormalities, whereas the UMLS integrates broader and richer information spanning drugs, disorders, procedures, laboratory tests, devices, anatomy, genes, proteins and more that can be used to capture non-phenotype signals for more robust RDR. Second, UMLS concepts aggregate extensive synonyms, abbreviations, and lexical variations that commonly appear in clinical texts, and thus improve the ability to match diverse textual expressions of the same concept across heterogeneous real-world clinical documentation. The extracted UMLS concepts, together with the LLM-produced disease profiles from which the concepts were derived, constitute the RDK and are organized into a unified knowledge base, PheMAP-RD, allowing a direct mapping between clinical narratives and machine-actionable concepts. While PheMAP-RD focuses on disease-level, concept-centric knowledge, more contextual signals are considered in our downstream pipelines. To preserve provenance and support source-specific assessment, knowledge from different sources (a specific LLM or NORD reports) is maintained and evaluated separately.

The multi-faceted RDK quality evaluation combines automated quality checks with targeted expert review. Automated metrics capture key properties of the generation process and the resulting RDK, including execution time, token usage, readability, reference composition, and clinical concept coverage. These analyses are complemented by clinical expert review (V.E.K. and M.G.) of the produced RDK layer for multiple randomly selected rare diseases. This evaluation framework is intended to be applied at each release of disease profiles, with automated checks run for every update and expert review used for quality assurance on sampled diseases and high-impact changes. These expert reviews are part of GEN-KnowRD’s evaluation process and thus are distinct from the expert curation of NORD reports. See Methods for details.

GEN-KnowRD supports general-purpose rare disease screening by introducing a novel two-stage pipeline that ranks the most likely candidate diseases based on a patient’s clinical presentation description (e.g., a summarized document or raw clinical notes). In the first stage, the clinical presentation of a patient first undergoes the same UMLS concept extraction process as mentioned earlier, such that a sparse representation is constructed in the form of a discrete UMLS concept list. GEN-KnowRD then applies the BM25 algorithm^45^ to calculate the similarity between this sparse representation and that of each rare disease profile, resulting in a concept-based ranked list of rare diseases. Rather than solely relying on sparse representations that focuses on lexical overlap, GEN-KnowRD also calculates semantic similarity using embedding-based dense representations to generate an independent ranked list of diseases. We selected and finetuned Qwen3-Embedding-8B^46^ for embedding generation due to its superior performance (Supplementary **Table 2**). We then used reciprocal rank fusion^47^ to aggregate sparse and dense disease rankings to derive a fused ranking score at the end of the first stage. The second stage starts with the top 20 diseases according to the fused scores, where each candidate disease (in the form of the combination of disease name and selected sections in the corresponding disease profile) is re-evaluated against the patient’s clinical presentation description using a reranker (Qwen3-reranker-8B^46^) to derive an updated disease ranking.

GEN-KnowRD can also perform specialized rare disease discrimination, where the goal is to evaluate a single candidate diagnosis. Given the clinical presentation description of a patient and a hypothesized diagnosis, a task-specific model determines whether the presentation is consistent with that diagnosis. To facilitate early identification of idiopathic pulmonary fibrosis (IPF) as a use case, a condition known for prolonged symptom-to-diagnosis delays^48,49^, we designed two diagnostic classification studies: 1) distinguishing IPF from non-IPF pulmonary diseases that may present with similar nonspecific symptoms and are often documented earlier in the diagnostic journey, and 2) distinguishing IPF from suspected IPF cases, targeting a clinically important pre-clinical suspicion window of high diagnostic uncertainty when accurately discriminating true IPF among suspected cases can accelerate referral and enable timely entry into IPF-focused care pathways. Using the established IPF RDK, GEN-KnowRD extracts key temporal features from patients’ longitudinal clinical notes and fits classifiers to adjudicate the diagnosis.

To evaluate the utility of LLM-generated rare disease profiles, as well as the performance of the supported general-purpose disease screening pipeline, we assembled six public rare disease benchmarks (**Fig. 1b**), including 1) HMS^50^, 2) LIRICAL^51^, 3) MME^52^, 4) RAMEDIS^53^, 5) MyGene2^54^, and 6) PMC-Patients^55^. We combined the first five benchmarks into a single one (Non-PMC) and converted the HPO terms used to describe patient clinical presentation into free-text descriptions. The PMC benchmark (i.e., PMC-Patients; free-text patient summaries) accounts for 92.6% (8,601/9,290) of patients and includes 651 unique rare diseases, whereas Non-PMC contains 7.4% (689/9,290) of patients and 40 unique rare diseases (**Fig. 1b,c**). The two benchmarks share 107 overlapping diseases (**Fig. 1d**). Disease complexity, defined as the number of Orphanet classification assignments (31 categories in total) according to affected body systems^56^, exhibits a long-tailed distribution, with most diseases assigned to relatively small complexity (**Fig. 1e**). We also constructed two real-world private diagnostic benchmarks (511 patients in total; Supplementary **Table 3**) based on longitudinal electronic health records (EHR) data from Vanderbilt University Medical Center for specialized rare disease discrimination (**Fig. 1f**). Detailed dataset preprocessing information is provided in the Methods.

We evaluated three baseline approaches for comparison. The first is the NORD rare disease reports^38^, which serves as a gold-standard knowledge source. We compared their utility against LLM-generated rare disease profiles for supporting RDR while keeping the GEN-KnowRD’s RDR pipelines fixed. The second baseline is PhenoBrain^43^, a state-of-the-art, HPO-first RDR framework that reflects the prevailing two-stage RDR paradigm. This approach first maps a patient’s clinical presentation description into HPO terms and then ranks diseases using an ensemble model grounded in human-curated knowledge. Third, we evaluated GPT-5 as a representative advanced LLM baseline for RDR to assess whether GEN-KnowRD, a lightweight framework, can achieve performance comparable to substantially more computationally intensive end-to-end LLM reasoning approaches.

### Quality of generative-AI based RDK

Rare disease profile generation demonstrates distinct patterns across LLMs (**Table 1**). First, OpenAI o3 and Gemini 2.5 Pro show the shortest prompt-to-response times, generating each disease profile in approximately 88 seconds, whereas Claude Sonnet 4 and DeepSeek R1 require significantly longer amounts of time (more than 140 seconds per disease). Second, Claude Sonnet 4 consumes the largest prompt contexts (323,766 tokens per disease), due to external web search and the integration of extensive retrieved information; it also generates the most verbose rare disease profiles (5,230 tokens per disease). OpenAI o3 ranks second for both input and output token usage. By contrast, Gemini 2.5 Pro and DeepSeek R1 consume input contexts that are roughly two orders of magnitude smaller and output shorter disease profiles. Third, other than Claude Sonnet 4, whose reasoning information is not available, OpenAI o3 triggers the reasoning function the most (37.5%) and leads to more reasoning tokens per disease (1,288), followed by Gemini 2.5 Pro and DeepSeek R1.

We observe multiple notable semantic characteristics in the rare disease profiles that were produced. Readability scores for all LLM-generated disease profiles, as well as the NORD reports, fall within the college-to-graduate range based on the Simple Measure of Gobbledygook metric (SMOG) (**Fig. 2a**), consistent with our prompt’s intent that the summaries “serve as a reliable Wikipedia entry for use by medical researchers and healthcare professionals”. Among the models, Gemini 2.5 Pro produces the most readable texts (corresponding to the lowest readability score), whereas OpenAI o3 shows the highest linguistic complexity and thus requires greater reading capability. Claude Sonnet 4 and DeepSeek R1 yield readability levels that are the most similar to the expert-curated reports. Since LLMs employ different evidence retrieval strategies, they exhibit distinct citation patterns. Gemini 2.5 Pro and Claude Sonnet 4 produce roughly 20 references per disease, whereas OpenAI o3 generates about 14 (**Fig. 2b**). Given that DeepSeek R1 does not support web search through an application programming interface (API) at the time of our experiments, it does not produce any references. Across models, references are most frequently drawn from government or official biomedical databases, such as PubMed, PMC, and NCBI, which index much of the authoritative peer-reviewed literature (**Fig. 2c**). The LLMs also rely heavily on specialized rare disease databases like Orphanet, which offer curated, high-specificity information about rare disorders. In addition, clinical or medical reference (e.g., Medscape, Cleveland Clinic Health Library, and Mayo Clinic Diseases and Conditions) and encyclopedia (e.g., Wikipedia) platforms are commonly cited for practical details on disease symptoms, diagnosis, and treatment guidelines. Academic journals or research portals, such as ScienceDirect, Wiley Online Library, MDPI, and ResearchGate, also contribute a nontrivial portion of citation coverage. Nonetheless, the usage of these three information categories is more variable across LLMs compared to government or official biomedical databases. Overall, Claude Sonnet 4 shows wider citation coverage than other models.

At the concept level, we observe that the number of extracted unique UMLS concepts varies across clinically relevant semantic groups and across LLMs (**Fig. 2d**). The “symptoms & conditions” group contributes the largest share of matches, followed by “genetics & molecular biology”, which is consistent with the goal of orienting disease profiles to support RDR. By contrast, the “drugs & procedures” and “diagnostics & laboratory findings” groups contain far fewer concepts across models, reflecting the lack of effective treatments and diagnostic approaches for rare diseases. Across model outputs, the disease profiles produced by Claude Sonnet 4 match the largest number of distinct UMLS concepts in all semantic groups, suggesting greater capability for evidence retrieval in RDR; OpenAI o3 and DeepSeek R1 consistently rank second and third. In comparison, the expert-curated NORD reports and Gemini 2.5 Pro match the fewest UMLS concepts across groups. Group-level similarity analysis suggests that UMLS concepts in “symptoms & conditions” are the most consistent among all groups, whereas “genetics & molecular biology” shows the greatest divergence (**Fig. 2e**). On average, the NORD reports exhibit lower similarity to LLM-produced profiles across all groups than the similarity observed among LLM-produced profiles themselves. Taken together with their smaller counts in unique UMLS concepts, this pattern suggests a narrower scope than the broader generative coverage produced by LLMs, which may enrich or extend the conceptual space beyond expert-curated content.

Two clinical experts independently assessed profiles for four randomly selected diseases (Behcet’s syndrome, granulomatosis with polyangiitis, IPF, and myasthenia gravis) across four dimensions: factual accuracy, clinical completeness, utility, and specificity. Inter-rater agreement measured with Cohen’s Kappa is substantial (0.644), according to Landis and Koch criteria.^57^ Across these diseases, LLM-generated profiles consistently exceed the expert-curated NORD reports in most dimensions (**Fig. 2f**). NORD reports rank lowest in clinical completeness, utility and disease specificity, and lower than DeepSeek R1- and Gemini 2.5 Pro-generated profiles in factual accuracy. Expert reviewers attributed specific shortcomings in the NORD IPF report to reliance on outdated clinical guidelines and conflation of drug-induced and radiation-induced pulmonary fibrosis with IPF. Among LLM-generated profiles, DeepSeek R1 and OpenAI O3 receive the highest ratings for clinical completeness, utility and disease specificity, while DeepSeek R1 and Gemini 2.5 Pro rank highest in factual accuracy. Claude Sonnet 4 ranks in the middle tier across most dimensions. Expert reviewers noted that Claude Sonnet 4-generated IPF profile references clinically unavailable diagnostic test (e.g., BAL SP-A levels, genetic testing) as though they play a routine role in diagnosis. These findings suggest that, at least for the diseases evaluated, LLM-generated profiles can achieve clinician-assessed quality that surpasses or equals established expert-curated resources, while also revealing dimension-specific variation across models. Interestingly, these expert ratings do not directly predict downstream computational performance as shown in the following sections.

### General-purpose rare disease screening evaluation

There are several notable findings for the general-purpose rare disease screening using GEN-KnowRD. First, across all knowledge sources, Stage 2 reranking uniformly refines Stage 1 disease rankings, improving Recall@1, @3, and @5 (**Fig. 3a**). For example, with expert-curated NORD rare disease reports, Stage 2 yields a Recall@1 of 0.837, which surpasses Stage 1’s rankings (0.613) by 36.5%. Second, LLM-produced rare disease profiles achieve consistently better RDR performance across both stages compared to expert-curated reports. Notably, Claude Sonnet 4 achieves a Stage 1 Recall@1 of 0.690, corresponding to a 12.6% increase over NORD reports (0.613). This advantage persists in Stage 2, although the improvement reduces to 2.7%. Third, Claude Sonnet 4 outperforms other LLMs on Recall@1, and, together with OpenAI o3, achieves the highest Recall@3 and Recall@5. This suggests that the rare disease profiles generated by Claude Sonnet 4 provide stronger support for RDR. Fourth, a Stage 2 variant that reranks diseases using only the disease names (without incorporating knowledge) performs uniformly worse than standard Stage 2 across all LLMs, highlighting the value of Stage 2’s knowledge-boosted design. Fifth, mean reciprocal rank (MRR), calculated as the average of the reciprocal ranks of the ground-truth diseases (with a range from 0 to 1), is leveraged to summarize the overall ranking quality for each knowledge source **(Fig. 3b)**. The LLM-produced knowledge layer (i.e., disease profiles) consistently outperforms NORD reports, yielding statistically significantly higher MRR across Stages 1 and 2. Among these sources, Claude Sonnet 4 performs best in Stage 1, whereas Claude Sonnet 4 and OpenAI o3 both lead in Stage 2.

**Fig. 3:**
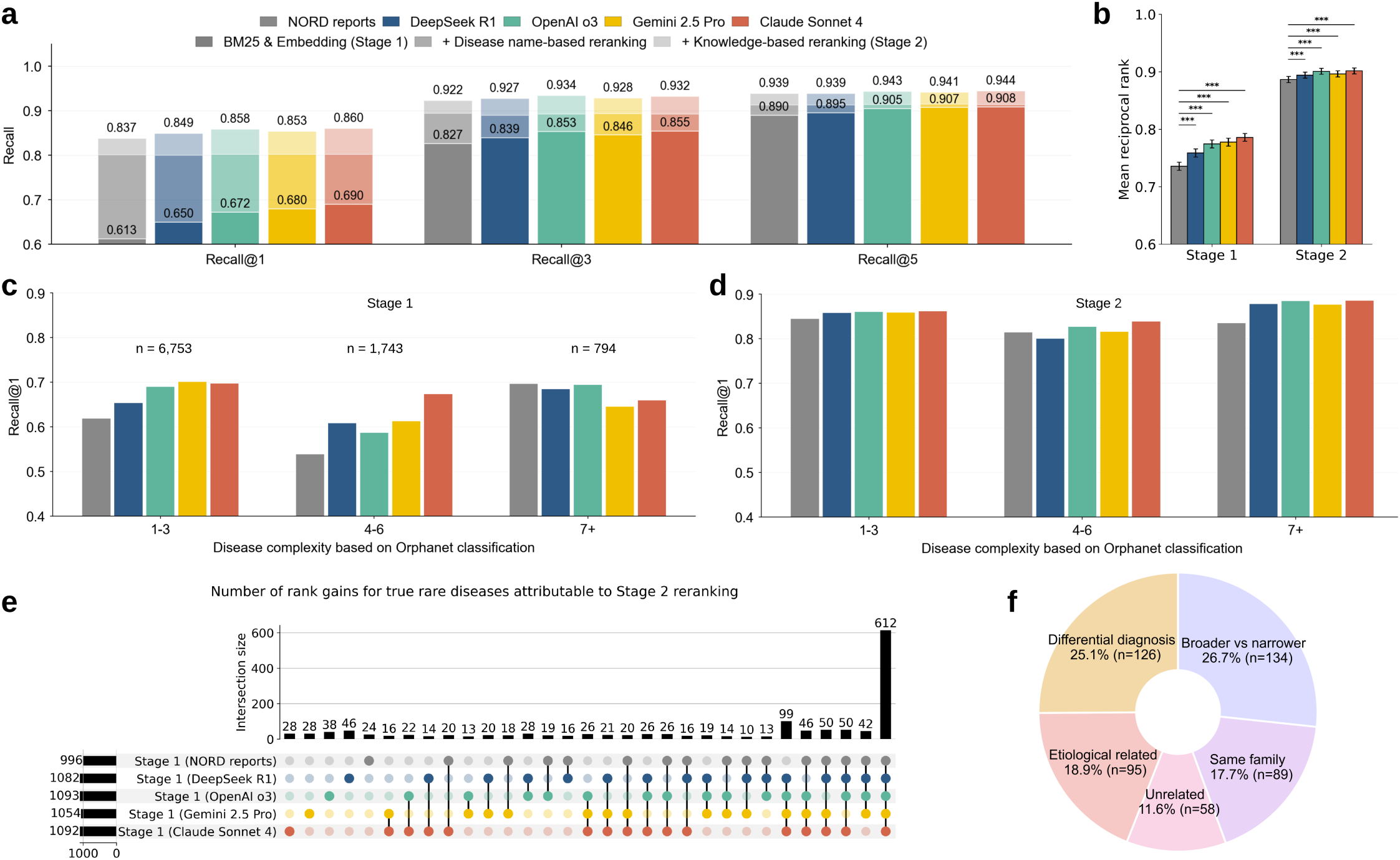
Stage-wise evaluation of GEN-KnowRD’s general-purpose rare disease screening. **a,** Recall for Stage 1 and Stage 2 across knowledge sources, along with a Stage 2 variant that reranks diseases using only disease names. **b,** Distributions of the mean reciprocal rank (MRR) of the ground-truth disease across knowledge sources. MRR differences between each LLM-based knowledge source and the NORD reports were evaluated using two-sided Wilcoxon signed-rank tests. **c,** Recall@1 of disease rankings out of Stage 1 across disease complexity categories based on Orphanet classification. The number of patents within each group is indicated. **d,** Recall@1 of disease rankings out of Stage 2 across disease complexity categories based on Orphanet classification. **e,** UpSet plot showing the number of rank gains of the ground-truth rare diseases attributable to knowledge-boosted reranking in Stage 2. The leftmost bar indicates the number of patients for whom the ground-truth disease moves up from Stage 1 position. **f,** Relationship categories identified in cases where Claude Sonnet 4 ranks the ground-truth rare disease second. *** indicates p<0.001 with Holm-Bonferroni correction.

We further evaluated GEN-KnowRD by disease complexity and observe heterogeneity in RDR performance **(Fig. 3c,d)**. The variability within and across complexity levels is more pronounced in Stage 1 than in Stage 2, consistent with the observed recall patterns above. At Stage 1, GEN-KnowRD, on average, exhibits a lower performance for diseases affecting four (Recall@1=0.624 across knowledge sources), five (0.600), or six (0.568) body systems (overall mean Recall@1=0.597), relative to those with fewer affected systems (overall mean Recall@1=0.672) or more systems (overall mean Recall@1=0.691). A similar phenomenon persists in Stage 2, although the variability is largely attenuated by the knowledge-boosted reranking. Across knowledge sources, Claude Sonnet 4-generated disease profiles achieve the strongest results, ranking best in six of nine complexity levels at Stage 1 and five out of nine at Stage 2 (Supplementary **Fig. 3**). By contrast, NORD reports often perform worst (seven of nine levels at Stage 1 and five of nine at Stage 2).

The benefit of knowledge-boosted reranking (Stage 2) is further reflected in rank gains for ground-truth diagnoses (**Fig. 3e**). Across knowledge sources, approximately 1,000 patients have their Stage 1 ranks of the true diagnoses improved, highlighting the reranker’s ability to leverage nuanced clinical evidence when aligning patient presentation with disease knowledge. Of these improvements, about 60% (612 patients) are shared across all knowledge sources, representing the largest subset. The second largest subset of rank gains is shared by all knowledge sources from LLMs (99 patients), whereas improvements involving the NORD reports occur in substantially smaller subsets, each comprising no more than half of that size.

Beyond aggregate RDR performance, we performed a ranking discrepancy analysis using GEN-KnowRD with the Claude Sonnet 4-produced disease profiles as an example (**Fig. 3f**). We used three LLMs as judges (Gemma-3-27B, OpenAI GPT-5, and OpenEvidence) and took their consensus to characterize the relationship between the top-rank disease and the true diagnosis. Among the 502 patients for whom the true disease is ranked second (accounting for approximately 80% of top-3 ranking discrepancies), the most common pattern is a broader-narrower relationship, where one disease is a parent entity or subtype of the other, accounting for 26.7% of the discrepancies. The remaining discrepancies are attributed to differential diagnosis pairs that are commonly confused clinically (25.1%), etiologic relationships where one can cause or lead to the other (18.9%), diseases within the same family (17.7%), and unrelated pairs with no meaningful links (11.6%). See detailed process in Methods.

Beyond evaluating a single knowledge source for RDR, we further examined the feasibility of leveraging all LLM-produced knowledge sources jointly. Specifically, we explored a gated ensemble strategy (GEN-KnowRD-Ensemble) that considers rankings from GEN-KnowRD instantiated with different LLM-produced knowledge sources, and compared it with two state-of-the-art RDR approaches (i.e., PhenoBrain and OpenAI GPT-5), as well as with GEN-KnowRD using the NORD reports. Among 7,038 patient cases with valid outputs from all approaches, GEN-KnowRD-Ensemble outperforms OpenAI GPT-5 with statistical significance across Recall@1, Recall@2, and Recall@3 (**Fig. 4a-c**). One exception is Recall@1 on PMC (6,364 patients), where OpenAI GPT-5 shows a slight better performance. This might be attributable to the free-text nature of PMC and potential data leakage during model pretraining. In contrast, OpenAI GPT-5 performs significantly worse on Non-PMC (674 patients) across Recall@1, Recall@2, and Recall@3. For example, GEN-KnowRD-Ensemble achieves a Recall@1 improvement of 129.1% than OpenAI GPT-5. This result might be attributable to Non-PMC benchmarks encoding a patient’s clinical presentation as individual HPO identifiers, for which LLM pretraining is less likely to capture the meaningful associations between these terms and true diagnoses at the free-text level.

**Fig. 4:**
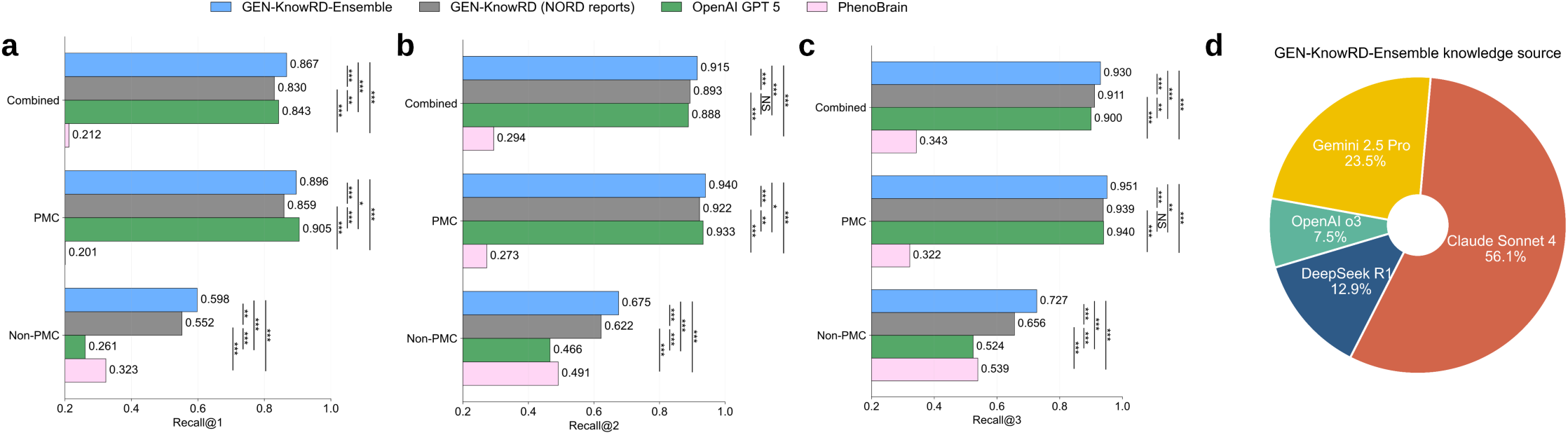
Evaluation of GEN-KnowRD-Ensemble against representative RDR approaches. **a,** Recall@1. **b,** Recall@2. **c,** Recall@3. **d,** Distribution of knowledge sources that GEN-KnowRD-Ensemble adopts in determining the final rare disease rankings. ***, **, and * indicate p<0.001, p<0.01, and p<0.05, respectively, from a two-sided Wilcoxon signed-rank test with Holm-Bonferroni correction. The prompt used for OpenAI GPT-5 is provided in Supplementary **Table 4**. NS: not significant.

Moreover, GEN-KnowRD-Ensemble also yields significantly higher recall than PhenoBrain across benchmarks, underscoring the limitations of pipelines that depend primarily on HPO extraction and mapping. For example, GEN-KnowRD-Ensemble achieves a Recall@1 gain of 345.8% relative to PhenoBrain on PMC. Nevertheless, PhenoBrain still outperforms OpenAI GPT-5 on Non-PMC, highlighting the value of compact, task-specialized models for RDR. Furthermore, GEN-KnowRD-Ensemble surpasses GEN-KnowRD with NORD reports, especially on Non-PMC (corresponding to a Recall@1 gain of 8.3%), demonstrating the added benefit of combining multiple LLM-derived knowledge sources through the gated ensemble mechanism. Detailed comparison results between GEN-KnowRD-Ensemble and GEN-KnowRD with individual knowledge sources are provided in Supplementary **Table 5**. Analysis of the knowledge source selected by the gating mechanism in GEN-KnowRD-Ensemble suggests that Claude Sonnet 4-derived knowledge source is used most frequently (56.1%), followed by Gemini 2.5 Pro (23.5%), DeepSeek R1 (12.9%), and OpenAI o3 (7.5%) (**Fig. 4d**).

### Specialized rare disease discrimination evaluation

We used IPF as a case study to evaluate GEN-KnowRD for specialized rare disease discrimination. In this setting, the number of extracted UMLS concepts from the IPF disease profiles varies across knowledge sources (**Fig. 5a**). The NORD IPF report contains 278 distinct UMLS concepts (the largest set), whereas OpenAI o3-generated IPF disease profile contains 154 concepts, representing the smallest set. Only 33 concepts overlap across all five sources, while 10 concepts are shared across the LLM-generated disease profiles, suggesting the strong uniqueness and diversity of knowledge representations across sources. Using the UMLS concepts extracted from each knowledge source, we identified their meaningful mentions in longitudinal clinical notes from the constructed cohorts in both early diagnosis studies (Supplementary **Fig. 4**) and then calculate, for each source, the difference in the total number of matched concepts between cases and controls. In Study 1 (IPF vs non-IPF), the disease profile generated by Claude Sonnet 4 leads to the largest case-control differences (**Fig. 5b**), indicating richer and more discriminative coverage of IPF-related characteristics, even though these sources do not have the highest number of distinct IPF concepts overall. By contrast, in Study 2 (IPF vs suspected IPF), the NORD IPF report corresponds to the largest case-control concept difference (**Fig. 5c**). Across both studies, OpenAI o3-generated IPF profile produces the smallest case-control concept differences.

**Fig. 5:**
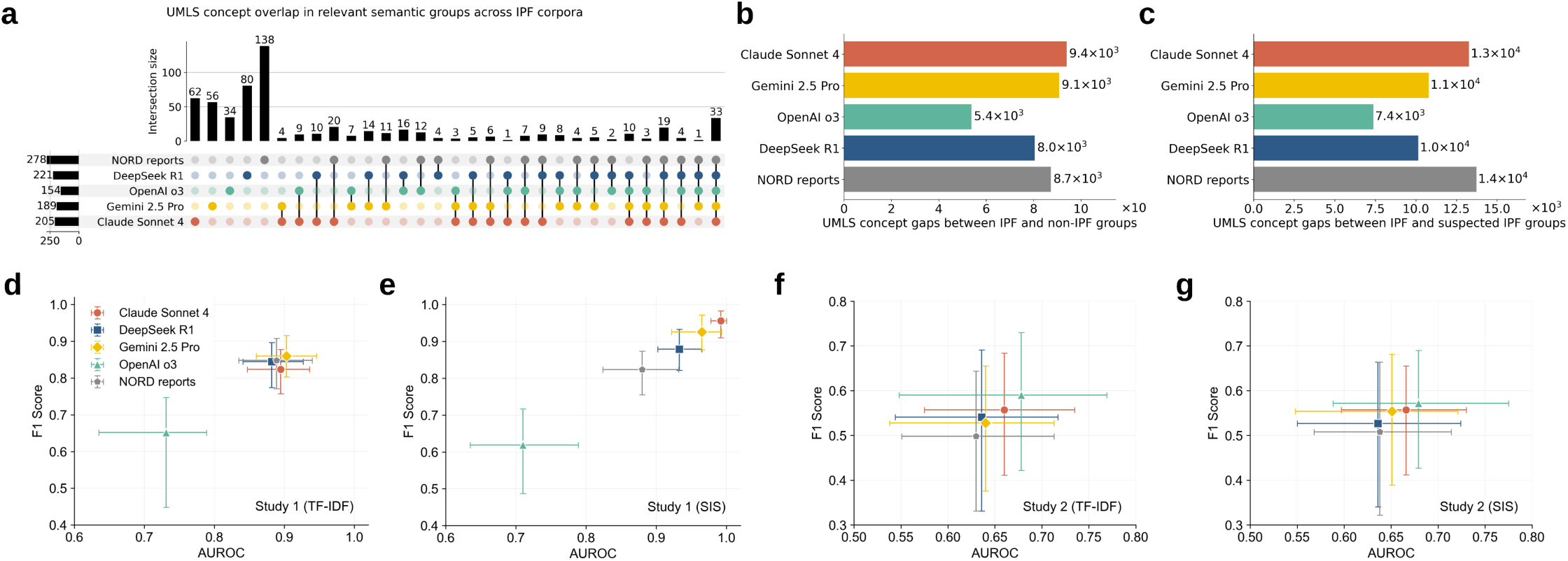
Evaluation of GEN-KnowRD in early diagnosis of idiopathic pulmonary fibrosis (IPF). **a,** UpSet plot showing overlap among UMLS concepts extracted from the clinical presentation sections of rare disease profiles. **b,** Differences in the total number of matched UMLS concepts extracted from longitudinal clinical notes of IPF group versus non-IPF group (169 patients per group; Study 1). Multiple mentions of the same concept within a single note are counted only once. **c,** Differences in the total number of matched UMLS concepts extracted from longitudinal clinical notes of IPF group versus suspected IPF group (171 patients per group; Study 2). **d,** AUROC and F1 scores of GEN-KnowRD with TF-IDF-weighted concept importance for early diagnosis of IPF in Study 1. **e,** AUROC and F1 scores of GEN-KnowRD with SIS-weighted concept importance for early diagnosis of IPF in Study 1. **f,** AUROC and F1 scores of GEN-KnowRD with TF-IDF-weighted concept importance for early diagnosis of IPF in Study 2. **g,** AUROC and F1 scores of GEN-KnowRD with SIS-weighted concept importance for early diagnosis of IPF in Study 2. Youden index is applied to determine cut-off thresholds and 95% confidence intervals for AUROC and F1 are presented for knowledge sources.

There are multiple notable findings in the performance of IPF early diagnosis. First, Studies 1 and 2 differ markedly in difficulty, as evidenced by substantially higher AUROC and F1 scores achieved in Study 1 than in Study 2 across all concept-weighting strategies (**Fig. 5d-g**). This difference is expected as IPF and other distinct pulmonary diseases tend to share fewer common clinical and physiological signatures in clinical notes than IPF and suspected IPF cases, whereas suspected IPF cases are enriched for fibrotic descriptors and diagnostic uncertainty, making the negative class highly confusable and the task inherently more difficult. Consequently, Study 2 requires resolving fine-grained, trajectory-dependent evidence beyond keywords, motivating more sophisticated approaches. Second, in Study 1, the OpenAI o3-produced IPF profile yields significantly lower performance than the NORD IPF report and other LLM-based sources, with AUROC of 0.731 (TF-IDF-weighted concept importance) and 0.710 (SIS-weighted) and F1 of 0.652 (TF-IDF) and 0.619 (SIS) (**Fig. 5d,e**). This result may be partially explained by the OpenAI o3-produced IPF clinical presentation lacking key terminology related to pulmonary fibrosis. In contrast, the corresponding AUROC and F1 scores for the NORD IPF report and other LLM-based sources are all above 0.800. Third, compared to TF-IDF-weighting, SIS-based weighting not only sharpens performance separation across knowledge sources, but also improves overall performance. Notably, the Claude Sonnet 4-based IPF profile achieves the highest AUROC (0.992) and F1 (0.956), corresponding to gains of 10.8% and 16.0% over the TF-IDF weighting setting, where most knowledge sources, except OpenAI o3, offer largely similar support for early diagnosis. Fourth, the differences of early diagnosis performance in Study 2 are more modest overall (**Fig. 5f,g**). Nevertheless, the NORD IPF profile shows relatively weaker performance than the LLM-produced disease profiles in terms of AUROC and F1. Interestingly, the average AUROC and F1 of OpenAI o3 rank highest in Study 2, suggesting that its symptom representations capture clinically meaningful distinctions within this challenging borderline population. The Claude Sonnet 4-generated IPF profile ranks second, achieving AUROC of 0.660 (TF-IDF) and 0.666 (SIS) and F1 of 0.557 for both weighting strategies.

## Discussion

The current limitations of RDR pose a major challenge that hinders timely intervention, cohort identification of patients for enrollment in clinical trials, and advances in therapeutic development. Yet current computational approaches for RDR depend on knowledge resources that are expensive to curate, structurally heterogeneous, incomplete, and slow to incorporate emerging evidence. To address these constraints at their source, we introduce GEN-KnowRD, a knowledge-first framework that generates schema-guided rare disease profiles, converts them into an AI-ready knowledge base (PheMAP-RD), and then links this resource with lightweight inference pipelines. Across six public benchmarks for general-purpose rare disease screening and two private longitudinal EHR cohorts for IPF early diagnosis, GEN-KnowRD consistently improves RDR relative to a state-of-the-art, HPO-centered baseline and to pipelines grounded in expert-curated NORD reports, while achieving stronger results than end-to-end LLM-based reasoning. These findings support our hypothesis that RDR is often constrained by how domain knowledge is presented and leveraged, and how clinical evidence is distilled into actionable signals, such that strengthening upstream knowledge layer and evidence processing can enable strong performance even with lightweight downstream strategies that can be deployed locally.

Our results imply broader translational value of GEN-KnowRD. Since PheMAP-RD provides structured, computable rare disease profiles that are decoupled from any single inference method, it can naturally function as shared infrastructure across clinical applications. For screening and triage, the general-purpose pipeline can flag patients whose records suggest unrecognized rare conditions, which potentially support earlier specialist referral and prompting targeted genetic testing. For clinical research, the same knowledge base can be repurposed to match patient phenotypic profiles against trial eligibility criteria, accelerating cohort identification for rare disease studies. Critically, the modular and reusable design of the knowledge layer means that PheMAP-RD is not limited to the pipelines presented in this paper. As end-to-end and agentic AI diagnostic systems continue to advance, exemplified by recent work such as DeepRare^28^, the quality of knowledge they retrieve and reason over will remain a key determinant of their reliability. GEN-KnowRD is designed to complement such systems by providing quality-assessed knowledge base that can strengthen evidence retrieval, grounding, and decision support. In this sense, GEN-KnowRD’s contribution is not a competing diagnostic tool, but a foundational layer that can raise the performance ceiling of the rare disease AI ecosystem.

Underneath the performance gains, the central contribution of GEN-KnowRD is a shift in how LLMs (and AI more broadly) are used in RDR. GEN-KnowRD employs LLMs to construct a disease knowledge layer that can be systematically evaluated, versioned, processed, and reused. This addresses multiple bottlenecks of the current knowledge-driven paradigm. First, by scaling the creation of disease profiles, it substantially reduces multi-disciplinary human curation effort required to construct and maintain rare disease knowledge resources. Traditional curation typically relies upon repeated manual synthesis by clinicians, geneticists, and clinical informaticians to perform literature review, reconcile fragmented evidence across publications and guidelines, normalize inconsistent terminology, and harmonize variable documentation practices across sources. In contrast, GEN-KnowRD shifts the workflow from labor-intensive end-to-end drafting to targeted review and governance, allowing scarce expert time to be focused on validation and auditing. This design also streamlines downstream processing since LLM-produced disease profiles become easier to evaluate, update, and integrate into AI-ready knowledge base that retrieval and reranking pipelines can consume directly, with fewer failures attributable to formatting issues or missing sections. Over time, this also enables faster refresh cycles, enabling a living resource in which updates can be applied systematically and consistently with minimal additional human effort.

Second, GEN-KnowRD strengthens the reliability and stability of knowledge-driven RDR by enforcing a shared schema and editorial constraints that make disease evidence easier to compare across conditions and less sensitive to source-specific idiosyncrasies. Compared to ad hoc, heterogeneous disease knowledge sources typically used for retrieval in end-to-end LLM workflows, this approach reduces representational bias and noise arising from uneven coverage, variable emphasis, and inconsistent terminology, leading to cleaner retrieval signals and more consistent disease prioritization. For example, in the heterogeneous knowledge setting, one disease may be described with detailed guideline-style sections, whereas another is captured in a short narrative that uses different synonyms and omits important information (e.g., testing cues). This can cause signal retrieval to disproportionately favor the better-structured disease simply because it contains more searchable anchors, rather than because it aligns more closely with the patient’s presentation.

Third, GEN-KnowRD enables controllable evidence pathways by separating knowledge processing from inference, which potentially supports the ability to diagnose and localize failures, as inaccurate prioritization or discrimination can be explicitly traced to specific stages (e.g., knowledge quality, concept extraction, retrieval, reranking, or feature selection), rather than being entangled within one big end-to-end, opaque reasoning process typically used by LLMs. Importantly, because inference is executed through fixed, lightweight components instead of interactive, per-query LLM generation, the behavior of GEN-KnowRD is more reproducible across runs and less vulnerable to prompt- or context-dependent variability that can otherwise complicate validation.

Fourth, compared to RDR workflows that directly invoke advanced proprietary LLMs and require transmitting sensitive patient health information to external services, GEN-KnowRD offers a more privacy-preserving and security-aligned deployment model. By shifting LLM use to an offline knowledge layer construction step and performing online inference via lightweight local computation, institutions can run the entire RDR workflow on-premise or within a secured private cloud, keeping protected health information inside their own network boundary. This minimizes reliance on third-party data handling and limits potential retention or secondary-use risks, while achieving tighter control than is typically feasible with external LLM calls.

Fifth, our design substantially cuts the computational and financial costs of RDR while achieving comparable or even greater performance than state-of-the-art approaches. GEN-KnowRD, along with PheMAP-RD, amortizes the expense of knowledge layer construction using LLMs across future patients and institutions, paving the way for scalable deployment without requiring per-query invocation of expensive LLM models or repeated long-context reasoning at inference time. These savings are especially pronounced in RDR based on EHR data, where longitudinal histories and clinical notes can be prohibitively lengthy to process directly with LLMs in routine workflows. Notably, PheMAP-RD is a durable community asset that can be freely reused by researchers and hospitals so that the primary cost is paid once during creation, while the benefits accumulate broadly through repeated downstream use.

In addition to these efficiency and scalability advantages, GEN-KnowRD also suggests a scalable pathway for maintaining a living rare disease knowledge layer, rather than a static resource that risks falling behind advances in clinical evidence. Because PheMAP-RD is built through a modular pipeline, it can refresh iteratively as stronger LLMs and new evidence become available. In an updated cycle, disease profiles can be regenerated or revised, clinical concept representation re-extracted, and the resulting knowledge base re-benchmarked before release. This design enables PheMAP-RD to evolve with medical knowledge and language modeling technology while lowering the maintenance burden relative to full manual re-curation. It also creates a practical path toward a community-facing rare disease knowledge resource that improves through transparent, versioned updates.

Beyond this forward-looking implication, the present study also reveals that LLM-constructed rare disease knowledge layer exhibits distinct characteristics that translate into measurable differences in RDR. Notably, they vary in reference patterns, linguistic complexity, and the breadth of clinical concepts captured across semantic groups. These differences reflect variations in LLMs’ retrieval strategies and synthesis style. Importantly, expert review indicates that human-curated NORD reports are not superior to LLM-produced disease profiles in key quality axes, including factual accuracy, clinical completeness, clinical utility, and clinical specificity. In fact, several LLM-generated disease profiles match or exceed NORD report on these criteria. However, we note that expert ratings are not expected to be fully consistent with downstream RDR performance. This is because RDR hinges not only on human perceived value but also on how well knowledge can be operationalized for computational matching, especially the coverage and discriminative value of extracted clinical concepts, as well as the extent to which retrieved evidence can be reliably aligned with heterogeneous clinical presentation of patients. The consistently strong performance of Claude Sonnet 4-produced disease profile across evaluation settings likely stems from their more comprehensive and clinically grounded representations, which in turn enable better evidence retrieval and disease ranking. Meanwhile, the diversity across LLMs motivates a source-level perspective that heterogeneous knowledge sources can be complementary, and the aggregation through gated ensemble can offer robustness beyond any single model’s knowledge synthesis capability.

Our results further suggest that RDR performance is influenced by how patient evidence is represented and linked to disease knowledge. Prior approaches that rely solely on HPO term extraction followed by phenotype matching based on the extracted terms (PhenoBrain as an example) inherit several structural disadvantages that become salient in real-world clinical narratives. First, HPO terms capture primarily phenotypic abnormalities and only partially reflect the breadth of diagnostically decisive evidence in rare diseases. Discarding non-phenotypic evidence and contextual information leads to a representation bottleneck that is difficult to overcome by supplicated matching strategies alone. Second, an HPO-first pipeline relies on accurate phenotype normalization from noisy text, yet clinical documentation is often ambiguous, contains negation and uncertainty, varies widely in granularity, and encodes important temporal information. All of these factors can induce systematic omissions or mis-mappings that propagate through the pipeline and irreversibly constrain the candidate disease set. Third, the phenotype-centered knowledge-matching can overemphasize the canonical presentations while underweighting atypical or evolving manifestations. When coupled with knowledge publication and curation biases in underlying resources, this can penalize ultra-rare diseases or evolving manifestations. Together, these factors explain why current HPO-first approaches may reach a performance ceiling in real-world RDR settings. In contrast, expanding the representation to UMLS concepts broadens the clinical signal space and improve robustness to lexical and documentation variation, while GEN-KnowRD’s lightweight downstream pipelines incorporate temporal and contextual cues during inference.

Consistent with this design rationale, our two-stage pipeline for general-purpose rare disease screening illustrates that knowledge-boosted reranking in Stage 2 consistently improves Recall@1, @2, and @5, compared to Stage 1 and brings performance to a more comparable range across knowledge sources. This convergence underscores the value of reranking as a knowledge-boosted refinement step. In particular, by re-evaluating the top-20 candidates with richer patient-disease interactions, reranking can recover signals that are only weakly captured by first-stage lexical or embedding similarity and mitigate source-specific noise in the initial retrieval rankings. Interestingly, reranking even without additional knowledge (e.g., using only disease names) can boost RDR performance to roughly the same level for all knowledge sources (**Fig. 3a**). This reflects the reranker’s (Qwen3-reranker-8B) ability to perform stronger cross-text alignment and better calibration within a small candidate set. Our observations are consistent with recent findings that highlight the value of reranking across various health applications^58–60^. We also note that the pretraining of the base model underlying the reranker might have been exposed to knowledge about diseases or medicine more broadly. Nonetheless, the consistently best results are achieved when reranking is guided by the core disease knowledge, underscoring the added value of reusable RDK beyond the name-only setting.

The ranking discrepancy analysis surfaces common weaknesses of current RDR approaches and assessment strategies and, at the same time, reveals future directions for improvement. The high prevalence of broader-narrower confusions suggests that GEN-KnowRD frequently identifies the correct diseases but cannot fully resolve fine-grained disease identity. These confusions may reflect issues in label granularity mismatches across catalogs and disease profiles that may emphasize shared manifestations more than discriminative subtype features. Addressing this limitation will require ontology-aware normalization, subtype-specific knowledge enrichment (e.g., adding more details that highlight what distinguishes a subtype from its parent), and refined evidence-aware reranking that prioritizes high-specificity cues. The large share of differential diagnosis and same-family discrepancies further reflects the RDR difficulty in settings where diseases share common symptoms and workups, arguably one of the biggest challenges for clinicians as well. Resolving these cases will require not only more comprehensive patient information collection but also higher-resolution modeling that captures temporality, severity trajectories, and discriminative evidence across similar diseases. Meanwhile, etiologically-related discrepancies suggest that the pipeline can prioritize diseases that are causally adjacent rather than the intended primary diagnosis, which might be addressed by exploiting causal directionality knowledge and explicitly representing the relationships between primary diseases and their comorbidities and complications.

Despite the merits of this work, our study has several limitations that should be acknowledged. First, we focused on approximately 1,300 diseases that have corresponding high-quality human-curated rare disease reports. Scaling GEN-KnowRD to the broader rare disease spectrum will require further validation of knowledge quality, consistency, and coverage, especially for ultra-rare and newly characterized conditions, and, critically, sufficiently rigorous labeled cohorts for benchmarking and evaluation. Second, although our baseline comparisons are conducted fairly, we ranked diseases within a rare disease candidate pool and did not explicitly include common conditions. This setting may simplify the real-world practice, where common diseases often compete strongly in the differential and can be confused with rare diseases due to shared manifestations. In future work, we intend to extend and rigorously evaluate GEN-KnowRD against the entire human disease catalog and enable it to function as an autonomous assistant for real-time early diagnosis across diverse clinical data environments, including EHRs. Third, LLM-generated disease profiles can vary with the choice of model, prompting strategy, retrieval behavior, and (when applicable) tool use, and the resulting knowledge may contain omissions or subtle inaccuracies. Although we conducted multi-faceted quality evaluation, fully characterizing provenance, timeliness, and factual reliability at scale remains challenging. At the same time, how best to optimize the RDK construction process and to maximize its downstream benefit for RDR are still open questions. Fourth, while GEN-KnowRD can acknowledge genotype and multimodal cues that can be expressed in text and mapped to UMLS concepts, the current pipelines, including ours, primarily operate on text-derived clinical evidence and concept-level representations. The incorporation of genetic variants, imaging findings, and laboratory patterns as first-class signals is likely to further improve both screening and discrimination, particularly for those defined by genotype-phenotype specificity or characteristic biomarker signatures. This may be addressed by integrating multi-modal foundation models to encode multi-modal information into latent representations and by coupling these embeddings with structured genotype features, which collectively enable joint retrieval and reranking over multi-modal evidence. Fifth, the two specialized IPF discrimination studies are derived from a single healthcare organization. As performance may vary across sites with different note writing practice, population characteristics, and care pathways, multi-center validation is needed to assess the transportability of our observations.

## Methods

### Dataset

#### NORD Rare Disease Database and Reports

The NORD Rare Disease Database compiles a comprehensive list of more than 10,000 rare diseases by aggregating information from established rare disease ontologies and reference sources like Orphanet and OMIM.^38^ From this extensive index, NORD content experts curate the full rare disease reports for a core subset of 1,320 diseases, each presenting summaries of clinical features, diagnosis, management, prognosis and additional resources. These reports are produced, reviewed, and maintained by medical experts who synthesize clinically vetted information from the medical literature, and they are freely available to support understanding, diagnosis, and care for rare diseases. We generated disease profiles for the 1,320 diseases that NORD has prioritized and use their reports as the primary knowledge baseline for comparison (**Fig. 1a**) given that they provide publicly accessible, clinically oriented, and well-organized information that is closely aligned with RDR using patients’ clinical notes or clinical presentation summaries. Other resources either are not uniformly designed as a clinically oriented description of diseases (e.g., OMIM) or vary substantially by content categories about a disease (e.g., Orphanet), making them less suitable to serve as a strong knowledge baseline.

#### Public Benchmarks

We use six public RDR benchmarks that have been widely referenced in prior research^28,29,43,61^: 1) HMS^50^, 2) LIRICAL^51^, 3) MME^52^, 4) RAMEDIS^53^, 5) MyGene2^54^, and 6) PMC-Patients^55^. PMC-Patients provides free-text patient summaries extracted from published case reports in PubMed Central, whereas the other five benchmarks are provided in a structured format consisting of HPO terms paired with ground-truth rare disease labels. We first mapped the disease labels of these benchmarks to the NORD rare disease report catalog through ORPHAcode and then converted HPO terms to their corresponding free-text descriptions, so that all input patient information in our experiments is represented as free text. For evaluation purposes, we combined the five benchmarks other than PMC-Patients into a single dataset (Non-PMC), and, for convenience, we refer to PMC-Patients as PMC (**Fig. 1a**). In total, 9,290 patients with 689 rare diseases are used in general-purpose disease screening. Note that PMC may not be suitable for evaluating early diagnosis, since some patients in the set have already been diagnosed. Instead, it serves as an appropriate benchmark for rare disease identification. Previous research has used LLMs to rewrite patient descriptions to remove diagnosis-related information to better align the input with the diagnostic task.^28^ However, this practice risk altering clinical meaning and potentially biasing the assessment of knowledge sources and leading to false conclusions. We therefore use the raw PMC data in our evaluation.

#### Private Benchmarks

In addition to the public benchmarks, we constructed a real-world early-diagnosis benchmark using raw, longitudinal clinical notes from patients at Vanderbilt University Medical Center. We designed two diagnostic tasks using IPF as an example to demonstrate 1) GEN-KnowRD’s capacity to discriminate rare diseases, and 2) the value of LLM-generated disease profiles. For the task of distinguishing IPF from non-IPF patients using data prior to their diagnosis, we define cases as individuals with not only preliminary ICD diagnosis codes but also billed ICD diagnosis codes (J84.112 or 516.31) documented in the EHR. We define controls as individuals with respiratory disease who received care from the same pulmonary specialists responsible for diagnosing IPF cases with no recorded IPF diagnosis codes. For each of the cases, we selected a control matched on sex, race, and age at disease onset within a ±5-year window. In addition, included patients must meet the following criteria: 1) disease onset age ≥45 years; 2) at least 90 days between the first documented visit and initial diagnosis; 3) at least 90 days between initial diagnosis and the last documented visit; 4) at least 30 clinical notes recorded between 18 and 3 months prior to diagnosis; and 5) at least 180-day spanning time of these notes within the pre-diagnosis window. For the task of distinguishing IPF from suspected IPF patients, we use the same definition of cases, while defining controls as individuals flagged in Vanderbilt University Medical Center’s clinical EHR database as only having preliminary IPF diagnosis codes but no corresponding billed IPF codes, using the database’s native indicators for preliminary versus billed diagnoses. Using these criteria, we identified 169 cases (and 169 matched controls) for IPF vs non-IPF comparison and 171 cases (and 171 matched controls) for IPF vs suspected IPF comparison. Detailed cohort construction is provided in Supplementary **Fig. 4.**

### Generation of RDK

#### RDK creation from generative AI models

We used Claude Sonnet 4, Gemini 2.5 Pro, OpenAI o3, and DeepSeek R1 to generate rare disease profiles with a prompt template approved by two clinical phenotyping experts (W.Q.W. and V.E.K.). We leveraged the corresponding application programming interfaces (API) for efficient data collection. To improve factual grounding and reduce hallucinated content and outdated information, we enabled the web search function in Claude Sonnet 4, Gemini 2.5 Pro, and OpenAI o3. In contrast, DeepSeek R1 did not provide a comparable capability through API during our experiments and thus was used without web search. To facilitate subsequent analysis, we employed Databricks MLflow Tracing to capture all details of the rare disease profile generation, including inputs, outputs, intermediate reasoning traces (if any), and metadata for each LLM invocation. For each rase disease and each LLM, a markdown file was created to store the corresponding disease profile.

#### Concept extraction

We applied SciSpacy^62^, a widely used open-source biomedical NLP toolkit, to extract relevant clinical concepts as it provides a practical, scalable pipeline with strong usability. We used its UMLS EntityLinker for concept extraction, with the similarity threshold set to 0.8. To identify concepts that are most relevant to the RDR tasks, we limited the concept extraction to four UMLS semantic groups: 1) symptoms & conditions, 2) diagnostics & laboratory findings, 3) drugs & procedures, and 4) genetics & molecular biology. The included semantic types under each group are summarized in Supplementary **Table 6**. We paired LLM-generated rare disease profiles with the corresponding extracted UMLS concepts to construct a rare disease knowledge base termed PheMAP-RD.

### Quality evaluation of generative-AI produced RDK

#### Objective metrics

Objective quality evaluation of rare disease profiles encompasses both the profile generation process and the properties of the produced profiles themselves. For the disease profile generation process for each LLM, we report running time, the number of input and output tokens, reasoning ratios, and external queries. Based on the generated disease profiles, we compare the following quantities across knowledge sources: 1) the categories and distributions of supporting references, 2) UMLS concepts matched across clinical semantic groups, and 3) text readability.

Readability is measured using the Simple Measure of Gobbledygook (SMOG)^63,64^ score, an established approach that estimates the education level required to understand a text. Specifically, SMOG calculates the number of polysyllabic words per 30 sentences and a higher value indicates more complex, less accessible writing, whereas lower scores reflect clearer and more readable content. In general, SMOG scores of 13-16 correspond to college-level readability, 17-18 to graduate-level, and 19+ to post-graduate or doctoral-level readability.^64^

#### Expert review

Two experts (V.E.K. and M.G.) with extensive experience in clinical phenotyping conducted quality evaluations of the collected rare disease profiles for four randomly selected diseases: Behcet’s syndrome, granulomatosis with polyangiitis, idiopathic pulmonary fibrosis, and myasthenia gravis. In addition to disease profiles produced by LLMs, the corresponding NORD rare disease reports were also evaluated using the same criteria. The source of each disease profile remains blind to review experts to minimize potential bias. The evaluation covers four major dimensions using a scale from 1 (worst) to 5 (best): 1) factual accuracy, which measures the degree to which statements, data, and claims in the disease profile are verifiable, correct, and consistent with current peer-reviewed evidence and established clinical guidelines, 2) clinical completeness, which measures the degree to which the disease profile covers essential clinical domains necessary for comprehensive understanding and management of the disease, without critical omissions, 3) clinical utility, which measures the degree to which the disease profile provides actionable, practically applicable information that supports clinical reasoning, decision-making, and patient care workflows, and 4) clinical specificity, which measures the degree to which the disease profile captures unique characteristics, distinguishing features, and context-specific considerations particular to the rare disease being described, avoiding generic or non-specific content. See Supplementary **Table 7** for detailed evaluation criteria.

Cohen’s Kappa with quadratic weights is leveraged to quantify inter-rater agreement, and the mean score across the two experts for each evaluation dimension is used to compare the quality across knowledge sources. We conducted dimension-wise error analysis based on the review documents collected from the review process.

### General-purpose rare disease screening

#### Stage 1: Sparse-dense similarity measurement

The overall goal of Stage 1 is to produce a short list of rare disease candidates that includes the ground-truth diagnosis. To generate an initial ranked list, we calculated the similarity between a patient’s clinical presentation descriptions and rare disease profiles using both sparse and dense representations. We use the BM25^45^ algorithm for sparse matching and embedding-based similarity for dense semantic matching. BM25 emphasizes exact term overlap and thus favors scenarios where disease specific terminology and explicit descriptors are present. In contrast, embedding-based similarity captures broader semantic relationships and can identify relevant diseases even when key terms are expressed differently or not stated verbatim. Because these methods excel under different linguistic conditions, combining them can leverage complementary signals and potentially yield more robust candidate prioritization than either method along.

We focus on four core sections of the generated rare disease profiles that capture the most clinically relevant information: 1) clinical presentation, 2) diagnostic evaluation, 3) subtypes or variants, and 4) management and standard therapy. Given that NORD rare disease reports do not consistently follow these section designs, we used an LLM to restructure the original reports into the same four categories without rewriting or adding new information. Specifically, we prompt Qwen3-235B-A22B to assign each sentence to one of the four sections (See the prompt in Supplementary **Table 8**).

Among the top three embedding models on the Massive Text Embedding Benchmark (MTEB) leaderboard^65^ as of September 2025, Qwen3-Embedding-8B^46^ and Llama-Embed-Nemotron-8B^66^ are open-source and suitable for further development. To determine their capability in real-world clinical retrieval settings, we constructed a disease prioritization dataset using 11 rare diseases (see the list in Supplementary **Fig. 5**) and clinical notes of patients from Vanderbilt University Medical Center spanning six months before to six months after first diagnosis. This dataset comprises 984 patient-disease pairs and 23,384 clinical notes, with note titles falling within: 1) progress notes, 2) assessment and plan, 3) subjective and objective, 4) admission history and physical exam, 5) discharge summary, 6) emergency provider notes, and 7) general clinic notes. We evaluated embedding models using semantic match rate (SMR), which is calculated in three steps: 1) for each rare disease candidate and each patient, we identify the patient’s clinical note with the highest embedding similarity to that disease, 2) we rank diseases based on the values of maximum similarity for the patient, and 3) we calculate the top-1 hit rate across patients. To simplify the embedding model selection, we first embedded only the name of rare disease, where Qwen3-Embedding-8B achieves a significantly higher top-1 SMR than Llama-Embed-Nemotron-8B (Supplementary **Table 2**). The embedding visualizations of clinical notes and disease names using t-SNE^67^ confirm the superiority of Qwen3-Embedding-8B (Supplementary **Fig. 5**). We then enriched disease representation by incorporating the four core sections described above, and further finetuned a Qwen3-Embedding-8B model for each LLM-generated disease profile using the same dataset through contrastive learning, where we constructed positive pairs using patients’ clinical notes and their corresponding diagnosed rare diseases and negative pairs through random selection. Because the finetuned Qwen3-Embedding-8B achieves the highest top-1 SMR (Supplementary **Table 2**), it is thus chosen as GEN-KnowRD’s embedding model. In this study, Qwen3-Embedding-8B was used to embed the full patient and disease descriptions without truncation.

To fuse the sparse and dense signals, we use reciprocal rank fusion (RRF) method^47^, a simple and robust strategy for aggregating ranked lists. The RRF-fused score of candidate disease *d* is defined as:

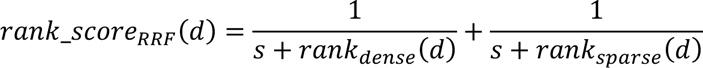

where *s* denotes a smoothing constant, set to 60 by following the original study^47^, and *rank_dense_* (*d*) and *rank_sparse_*(*d*) denote disease *d*’s positions in the dense and sparse rankings, respectively. Overall, RRF upweights diseases that appear near the top of either list and downweighs those that rank poorly in both, yielding a fused ranking score that is robust to noise and complementary failure modes across retrieval methods. We evaluated recall@5 to justify this fusion design of GEN-KnowRD, which outperforms either sparse-only or dense-only retrieval (Supplementary **Fig. 6**).

#### Stage 2: Rare disease reranking

To refine the disease rankings from Stage 1, which is built on the whole disease space, we applied a reranker to the top K diseases. Specifically, we use Qwen3-reranker-8B^46^ to re-evaluate pairwise relevance between a patient’s clinical presentation description and each of the K diseases. The inputs of the reranker are 1) a patient’s clinical presentation description, and 2) a disease document formed by combining the disease name with the core RDK sections. The reranker ingests the concatenated pair to align fine-grained clinical cues with disease-specific knowledge and then outputs a scalar relevance score reflecting how well the disease explains the patient’s clinical presentation. The relevance score is calculated independently for each of the K candidates, which are then sorted according to the calculated scores to form the final ranked list. Compared to the first stage that relies on lexical overlap and embedding similarity, the reranker in the second stage can exploit richer interaction features to upweight candidates supported by multiple concordant findings and downweigh those contradicted by explicitly negated evidence. Because we applied reranking only to a small candidate set (K=20), it remains computationally feasible while substantially improving precision at the top of the list.

#### Ensemble

As Stages 1 and 2 leverage the rare disease profiles produced by a single LLM, we further explored whether combining disease prioritizations from different LLMs can boost RDR performance. We implemented a Spearman rank correlation-based ensemble gating method that selects, for each patient, the most consensus-aligned disease ranking. Specifically, we computed the sum of pairwise rank correlations between each ranking and all others, and chose the ranking with the highest total agreement, aiming to reduce the chance of introducing spurious disease candidates from noisy lower-ranked predictions. We chose the top three diseases for correlation calculation since disease rankings beyond this threshold are more error-prone and less confident. We view this ensemble variant, GEN-KnowRD-Ensemble, as an aggregated pipeline for comparison with baseline models.

#### Discrepancy analysis

Since Claude Sonnet 4-produced rare disease profiles demonstrate the highest Recall@*x* for RDR compared to other LLMs and its performance gets saturated quickly from Recall@2, we examined all cases (n=502), where ground-truth diseases are ranked second by GEN-KnowRD, to summarize the relationship between those ranked first and the ground-truth diseases. We use Gemma-3-27B and GPT-5 to independently classify the relationships. When they output the same category, the relationship is accepted. When they identify different relationships, OpenEvidence is invoked to derive the final relationship. We define five relationship categories: 1) broader-narrower, where the disease ranked first (rank-1 disease) is a broader class or a specific subtype of the disease ranked second (rank-2 disease), 2) same-category, where both diseases belong to the same disease family or superclass, 3) etiologically related, where one disease may cause or lead to the other, 4) differential diagnosis, where the two diseases are commonly confused clinically, and 5) unrelated, where they involve different systems or etiologies with no meaningful links.

### Specialized rare disease discrimination

Clinical notes within the pre-diagnosis window (between 18 months and 3 months prior to diagnosis) from included patients first undergo the same UMLS concept extraction procedure as described above. Using the IPF disease profile generated by an LLM, the UMLS concepts appearing in its “clinical presentation” section are selected as the target set for clinical evidence extraction, as they capture clinically actionable, symptom-level descriptors most relevant to IPF diagnosis. To mitigate inaccurate evidence matching, we first applied MedGemma-27B^68^ to process all clinical notes and classify each concept as one of the following categories: family history, historical condition, hypothetical condition, negated condition, possible condition, or others. Only those classified as possible and others are used for following analysis. For each patient-week and each symptom concept, we assigned a three-level code, where 0, 1, and 2 indicate no mention of the symptom in that week’s notes, symptom mentioned that week, and symptom not mentioned despite other symptoms being mentioned in the same week, respectively. This results in a complete symptom-week matrix per patient, in which we identified all symptom concept pairs that co-occur within a week (i.e., both symptoms coded as 1). For each symptom pair, we compute two clinically motivated quantities: 1) the number of distinct co-occurrence weeks (*pair*_*week*_*single*) and 2) the number of co-occurrence weeks in which either symptom is labeled as worsening (*worsen*_*count*_*single*) by MedGemma-27B. These pair-level quantities are then aggregated to the patient level by summing across all symptom pairs to represent overall co-occurrence activity (*pair*_*week*_*agg*) and worsening burden ( *worsen*_*count*_*agg*). To weigh symptoms by discriminative relevance, each symptom concept *c* receives a symptom importance score (SIS) based on prevalence difference between IPF cases and controls, calculated as 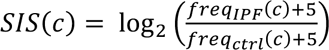, and a patient-specific symptom weight, calculated as *SIS*(*c*) ⋅ (1 + *freq_pa_(c)*), where *freq_pa_*(*c*) is the number of weeks in which *c* is present for that patient. We also calculate the term frequency-inverse document frequency (TF-IDF) value of concept *c* as an alternative importance score for comparison. To avoid double counting symptoms that appear in multiple co-occurring pairs, we pool all symptoms participating in any pair, deduplicated at the (patient, symptom) level, and sum the unique symptom weights to obtain total symptom weight (*total*_*symptom*_*weight*). Finally, we normalize overall symptom burden by co-occurrence activity using *normalized*_*weight* = *total*_*symptom*_*weight*/(*pair*_*week*_*agg* + 1). The downstream classifiers use *normalized*_*weight*, *pair*_*week*_*agg*, and *worsen*_*count*_*agg* as patient-level features to distinguish IPF cases from two different types of controls.

For each task, we fit a logistic regression model and evaluate model performance using stratified out-of-bag bootstrap validation^69^, with a 2:1 training-test ratio and 100 repetitions. Each trained classifier is evaluated on the held-out test set using standard measures that quantify both classification performance and the utility of the LLM-generated rare disease profiles for supporting early IPF diagnosis.

## Data Availability

LLM-generated rare disease profiles and the corresponding extracted UMLS concepts for each rare disease (i.e., PheMAP-RD) are available at https://wei-phenolib-api.app.vumc.org/. This service also hosts the patient clinical presentation descriptions (9,290) used in our public benchmark evaluations and GEN-KnowRD-Ensemble results. The private benchmarks for IPF early diagnosis are available upon request and IRB approval by Vanderbilt University Medical Center.

## Acknowledgements

This work is supported in part by National Institute of Health grants R01HG012748, K99LM014428, R00LM014429, R01HG013031, R01AG084550, R01HL171809, R01HG012748, R01LM012806, P50HD106446, R01GM139891, and UL1TR002243.

## Author Contribution Statement

W.Q.W., C.Y., and W.S. conceived and designed this study. W.S. and C.Y. conducted data preprocessing, performed the experiments, and analyzed the results. W.S. and Y.X. performed LLM data collection. M.E.G. and V.E.K. reviewed and evaluated LLM-generated rare disease profiles as clinical experts. C.Y. and W.S. summarized the major experimental findings and drafted the manuscript. W.Q.W., H.L., L.W., J.W., and R.L. assisted in interpreting the results and provided significant intellectual feedback. A.L.D., Q.F., C.S., V.A.B., J.L., C.M.S., K.W., P.J.E., and B.A.M. extensively revised the manuscript. W.Q.W. supervised the study. All authors participated in manuscript preparation and approved the final version.

## Ethics Declarations

There is no conflict of interest for this study.

## References

1. The Lancet Global Health. The landscape for rare diseases in 2024. Lancet Glob. Health 12, e341 (2024).

2. Baynam, G. et al. Global health for rare diseases through primary care. Lancet Glob. Health 12, e1192–e1199 (2024).

3. Taruscio, D. & Gahl, W. A. Rare diseases: Challenges and opportunities for research and public health. Nat. Rev. Dis. Primer 10, 13 (2024).

4. Faye, F. et al. Time to diagnosis and determinants of diagnostic delays of people living with a rare disease: Results of a Rare Barometer retrospective patient survey. Eur. J. Hum. Genet. 32, 1116–1126 (2024).

5. Schaefer, J., Lehne, M., Schepers, J., Prasser, F. & Thun, S. The use of machine learning in rare diseases: A scoping review. Orphanet J. Rare Dis. 15, 145 (2020).

6. Lagorce, D. et al. Phenotypic similarity-based approach for variant prioritization for unsolved rare disease: A preliminary methodological report. Eur. J. Hum. Genet. 32, 182–189 (2024).

7. Smail, C. et al. Complex trait associations in rare diseases and impacts on Mendelian variant interpretation. Nat. Commun. 15, 8196 (2024).

8. Chimirri, L. et al. Consistent performance of large language models in rare disease diagnosis across ten languages and 4917 cases. eBioMedicine 121, 105957 (2025).

9. Zhai, W., Huang, X., Shen, N. & Zhu, S. Phen2Disease: A phenotype-driven model for disease and gene prioritization by bidirectional maximum matching semantic similarities. Brief. Bioinform. 24, bbad172 (2023).

10. Jagadeesh, K. A. et al. Phrank measures phenotype sets similarity to greatly improve Mendelian diagnostic disease prioritization. Genet. Med. 21, 464–470 (2019).

11. Li, Q., Zhao, K., Bustamante, C. D., Ma, X. & Wong, W. H. Xrare: A machine learning method jointly modeling phenotypes and genetic evidence for rare disease diagnosis. Genet. Med. 21, 2126–2134 (2019).

12. Jia, J. et al. RDAD: A machine learning system to support phenotype-based rare disease diagnosis. Front. Genet. 9, 587 (2018).

13. Peng, J. et al. Measuring phenotype semantic similarity using Human Phenotype Ontology. in 2016 IEEE International Conference on Bioinformatics and Biomedicine (BIBM) 763–766 (IEEE, Shenzhen, China, 2016). doi:10.1109/BIBM.2016.7822617.

14. Zhao, M., et al. Phen2Gene: Rapid phenotype-driven gene prioritization for rare diseases. NAR Genomics Bioinforma. 2, lqaa032 (2020).

15. Pavan, S. et al. Clinical practice guidelines for rare diseases: The Orphanet database. PLOS ONE 12, e0170365 (2017).

16. Hamosh, A. Online Mendelian Inheritance in Man (OMIM), a knowledgebase of human genes and genetic disorders. Nucleic Acids Res. 33, D514–D517 (2004).

17. Landrum, M. J. et al. ClinVar: Public archive of interpretations of clinically relevant variants. Nucleic Acids Res. 44, D862–D868 (2016).

18. Shourick, J., Wack, M. & Jannot, A.-S. Assessing rare diseases prevalence using literature quantification. Orphanet J. Rare Dis. 16, 139 (2021).

19. Ibañez, K. et al. Increased frequency of repeat expansion mutations across different populations. Nat. Med. 30, 3357–3368 (2024).

20. Kariampuzha, W. Z. et al. Precision information extraction for rare disease epidemiology at scale. J. Transl. Med. 21, 157 (2023).

21. Nicholson, D. N., Himmelstein, D. S. & Greene, C. S. Expanding a database-derived biomedical knowledge graph via multi-relation extraction from biomedical abstracts. BioData Min. 15, 26 (2022).

22. Amberger, J. S., Bocchini, C. A., Schiettecatte, F., Scott, A. F. & Hamosh, A. OMIM.org: Online Mendelian Inheritance in Man (OMIM®), an online catalog of human genes and genetic disorders. Nucleic Acids Res. 43, D789–D798 (2015).

23. Vasilevsky, N. A. et al. Mondo: Integrating disease terminology across communities. GENETICS iyaf215 (2025).

24. Martínez-deMiguel, C., Segura-Bedmar, I., Chacón-Solano, E. & Guerrero-Aspizua, S. The RareDis corpus: A corpus annotated with rare diseases, their signs and symptoms. J. Biomed. Inform. 125, 103961 (2022).

25. Young, C. C. et al. Diagnostic accuracy of a custom large language model on rare pediatric disease case reports. Am. J. Med. Genet. A. 197, e63878 (2025).

26. Shyr, C. et al. Identifying and extracting rare diseases and their phenotypes with large language models. J. Healthc. Inform. Res. 8, 438–461 (2024).

27. Shyr, C. et al. Large language models for rare disease diagnosis at the undiagnosed diseases network. JAMA Netw. Open 8, e2528538 (2025).

28. Zhao, W. et al. An agentic system for rare disease diagnosis with traceable reasoning. Nature 10.1038/s41586-025-10097-9 (2026) doi:10.1038/s41586-025-10097-9.

29. Chen, X. et al. RareBench: Can LLMs Serve as Rare Diseases Specialists? In Proceedings of the 30th ACM SIGKDD Conference on Knowledge Discovery and Data Mining 4850–4861 (ACM, Barcelona Spain, 2024). doi:10.1145/3637528.3671576.

30. Pang, L., et al. Large language model sourcing: A survey. Preprint at 10.48550/arXiv.2510.10161 (2025).

31. Wang, H., Prasad, A., Stengel-Eskin, E. & Bansal, M. Retrieval-augmented generation with conflicting evidence. Preprint at 10.48550/ARXIV.2504.13079 (2025).

32. Masanneck, L., Meuth, S. G. & Pawlitzki, M. Evaluating base and retrieval augmented LLMs with document or online support for evidence based neurology. Npj Digit. Med. 8, 137 (2025).

33. Wang, S. et al. Knowledge editing for large language models: A survey. ACM Comput. Surv. 57, 1–37 (2025).

34. Yoran, O., Wolfson, T., Ram, O. & Berant, J. Making retrieval-augmented language models robust to irrelevant context. Preprint at 10.48550/arXiv.2310.01558 (2024).

35. Wu, K. et al. An automated framework for assessing how well LLMs cite relevant medical references. Nat. Commun. 16, 3615 (2025).

36. Rahman, S., et al. Generalization in healthcare AI: Evaluation of a clinical large language model. Preprint at 10.48550/arXiv.2402.10965 (2024).

37. Peng, Y. et al. From GPT to DeepSeek: Significant gaps remain in realizing AI in healthcare. J. Biomed. Inform. 163, 104791 (2025).

38. National Organization for Rare Disorders. Rare disease database. https://rarediseases.org/rare-diseases/ (2025).

39. Anthropic. Introducing Claude 4. https://www.anthropic.com/news/claude-4 https://www.anthropic.com/news/claude-4 (2025).

40. DeepSeek-AI et al. DeepSeek-R1: Incentivizing reasoning capability in LLMs via reinforcement learning. Preprint at 10.48550/ARXIV.2501.12948 (2025).

41. Google DeepMind. Gemini 2.5: Our Most Intelligent AI Model. https://blog.google/innovation-and-ai/models-and-research/google-deepmind/gemini-model-thinking-updates-march-2025 / https://blog.google/innovation-and-ai/models-and-research/google-deepmind/gemini-model-thinking-updates-march-2025/ (2025).

42. OpenAI. Introducing OpenAI O3 and O4-Mini. https://openai.com/index/introducing-o3-and-o4-mini/ https://openai.com/index/introducing-o3-and-o4-mini/ (2025).

43. Mao, X. et al. A phenotype-based AI pipeline outperforms human experts in differentially diagnosing rare diseases using EHRs. Npj Digit. Med. 8, 68 (2025).

44. PheMAP-RD. PheMAP-RD. https://wei-phenolib-api.app.vumc.org/ (2026).

45. Robertson, S. & Zaragoza, H. The Probabilistic relevance framework: BM25 and beyond. Found. Trends® Inf. Retr. 3, 333–389 (2009).

46. Zhang, Y., et al. Qwen3 embedding: Advancing text embedding and reranking through foundation models. Preprint at 10.48550/ARXIV.2506.05176 (2025).

47. Cormack, G. V., Clarke, C. L. A. & Buettcher, S. Reciprocal rank fusion outperforms condorcet and individual rank learning methods. In Proceedings of the 32nd international ACM SIGIR conference on Research and development in information retrieval 758–759 (ACM, Boston MA USA, 2009). doi:10.1145/1571941.1572114.

48. Grant-Orser, A. et al. The diagnostic pathway for patients with interstitial lung disease: A mixed-methods study of patients and physicians. BMJ Open Respir. Res. 11, e002333 (2024).

49. Onishchenko, D. et al. Screening for idiopathic pulmonary fibrosis using comorbidity signatures in electronic health records. Nat. Med. 28, 2107–2116 (2022).

50. Ronicke, S. et al. Can a decision support system accelerate rare disease diagnosis? Evaluating the potential impact of Ada DX in a retrospective study. Orphanet J. Rare Dis. 14, 69 (2019).

51. Robinson, P. N. et al. Interpretable clinical genomics with a likelihood ratio paradigm. Am. J. Hum. Genet. 107, 403–417 (2020).

52. Philippakis, A. A. et al. The Matchmaker exchange: A platform for rare disease gene discovery. Hum. Mutat. 36, 915–921 (2015).

53. Topel, T., Scheible, D., Trefz, F. & Hofestadt, R. RAMEDIS: A comprehensive information system for variations and corresponding phenotypes of rare metabolic diseases. Hum. Mutat. 31, E1081–E1088 (2010).

54. University of Washington. MyGene2. https://mygene2.org/MyGene2/ (2026).

55. Zhao, Z., Jin, Q., Chen, F., Peng, T. & Yu, S. A large-scale dataset of patient summaries for retrieval-based clinical decision support systems. Sci. Data 10, 909 (2023).

56. Orphanet. Orphanet Disease Classification. https://www.orpha.net/en/disease/classification/heads (2025).

57. Landis, J. R. & Koch, G. G. The measurement of observer agreement for categorical data. Biometrics 33, 159 (1977).

58. Wang, X., Mercer, R. & Rudzicz, F. Multi-stage retrieve and re-rank model for automatic medical coding recommendation. In Proceedings of the 2024 Conference of the North American Chapter of the Association for Computational Linguistics: Human Language Technologies (*Volume* 1: *Long Papers*) 4881–4891 (Association for Computational Linguistics, Mexico City, Mexico, 2024). doi:10.18653/v1/2024.naacl-long.273.

59. Yang, R. et al. KG-Rank: Enhancing large language models for medical QA with knowledge graphs and ranking techniques. In Proceedings of the 23rd Workshop on Biomedical Natural Language Processing 155–166 (Association for Computational Linguistics, Bangkok, Thailand, 2024). doi:10.18653/v1/2024.bionlp-1.13.

60. Kusa, W., Mendoza, Ó. E., Knoth, P., Pasi, G. & Hanbury, A. Effective matching of patients to clinical trials using entity extraction and neural re-ranking. J. Biomed. Inform. 144, 104444 (2023).

61. Yang, T., et al. A specialized large language model for clinical reasoning and diagnosis in rare diseases. Preprint at 10.48550/ARXIV.2511.14638 (2025).

62. Neumann, M., King, D., Beltagy, I. & Ammar, W. ScispaCy: Fast and robust models for biomedical natural language processing. In Proceedings of the 18th BioNLP Workshop and Shared Task 319–327 (Association for Computational Linguistics, Florence, Italy, 2019). doi:10.18653/v1/W19-5034.

63. Harvard Center for Health Communication. SMOG readability formula: Your tool for clearer health communication. https://hsph.harvard.edu/research/healthcommunication/resources/smog/?utm_source=chatgpt.com (2025).

64. Laughlin, G. H. M. SMOG grading-a new readability formula. J. Read. 12, 639–648 (1969).

65. MTEB Leaderboard. https://huggingface.co/spaces/mteb/leaderboard (2025).

66. Babakhin, Y., et al. Llama-Embed-Nemotron-8B: A universal text embedding model for multilingual and cross-lingual tasks. Preprint at 10.48550/ARXIV.2511.07025 (2025).

67. Maaten, L. van der & Hinton, G. Visualizing data using t-SNE. J. Mach. Learn. Res. 9, 2579–2605 (2008).

68. Sellergren, A., et al. MedGemma technical report. Preprint at 10.48550/ARXIV.2507.05201 (2025).

69. Fernandez-Felix, B. M., García-Esquinas, E., Muriel, A., Royuela, A. & Zamora, J. Bootstrap internal validation command for predictive logistic regression models. Stata J. Promot. Commun. Stat. Stata 21, 498–509 (2021).

