## Supplementary Data for "GEN-KnowRD: Reframing AI for Rare Disease Recognition"

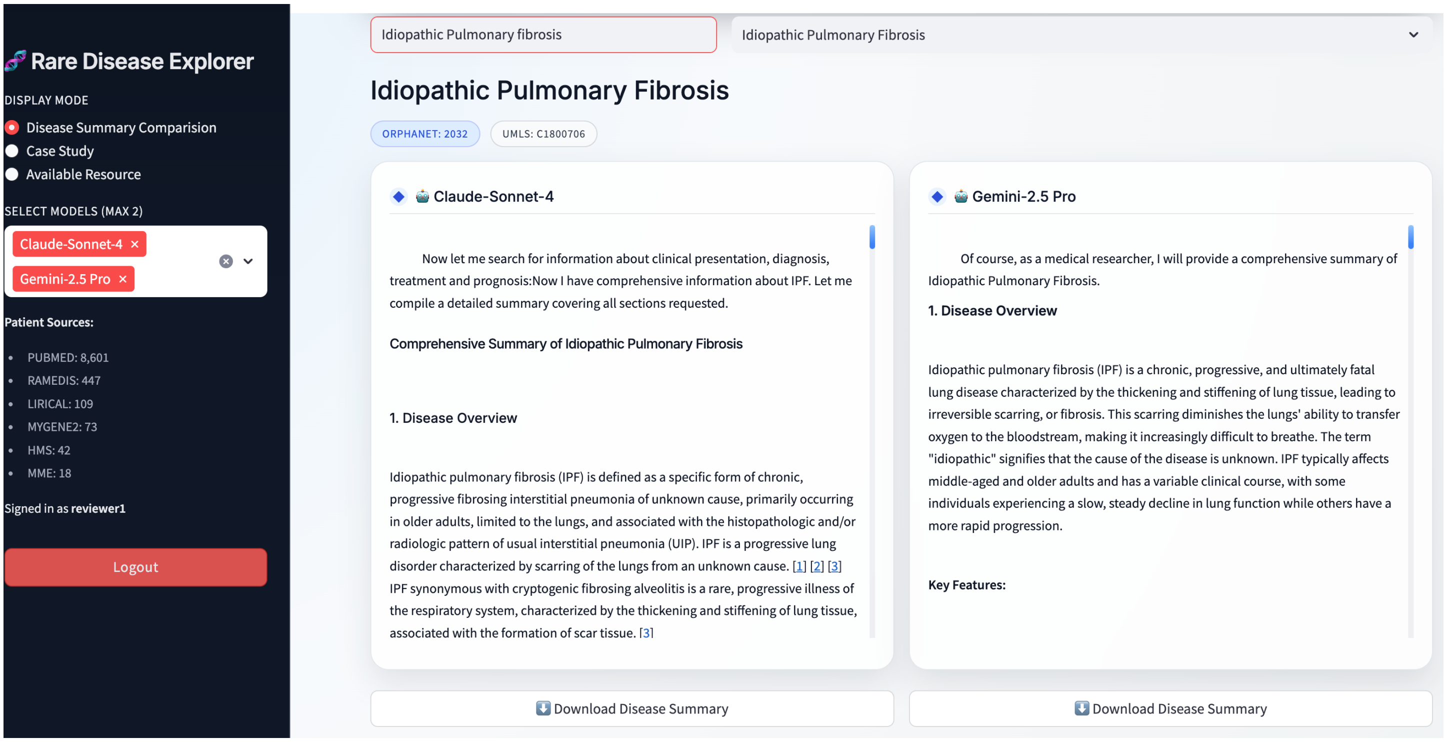


**Supplementary Fig. 1: Screenshot of large language model-produced rare disease profiles.**

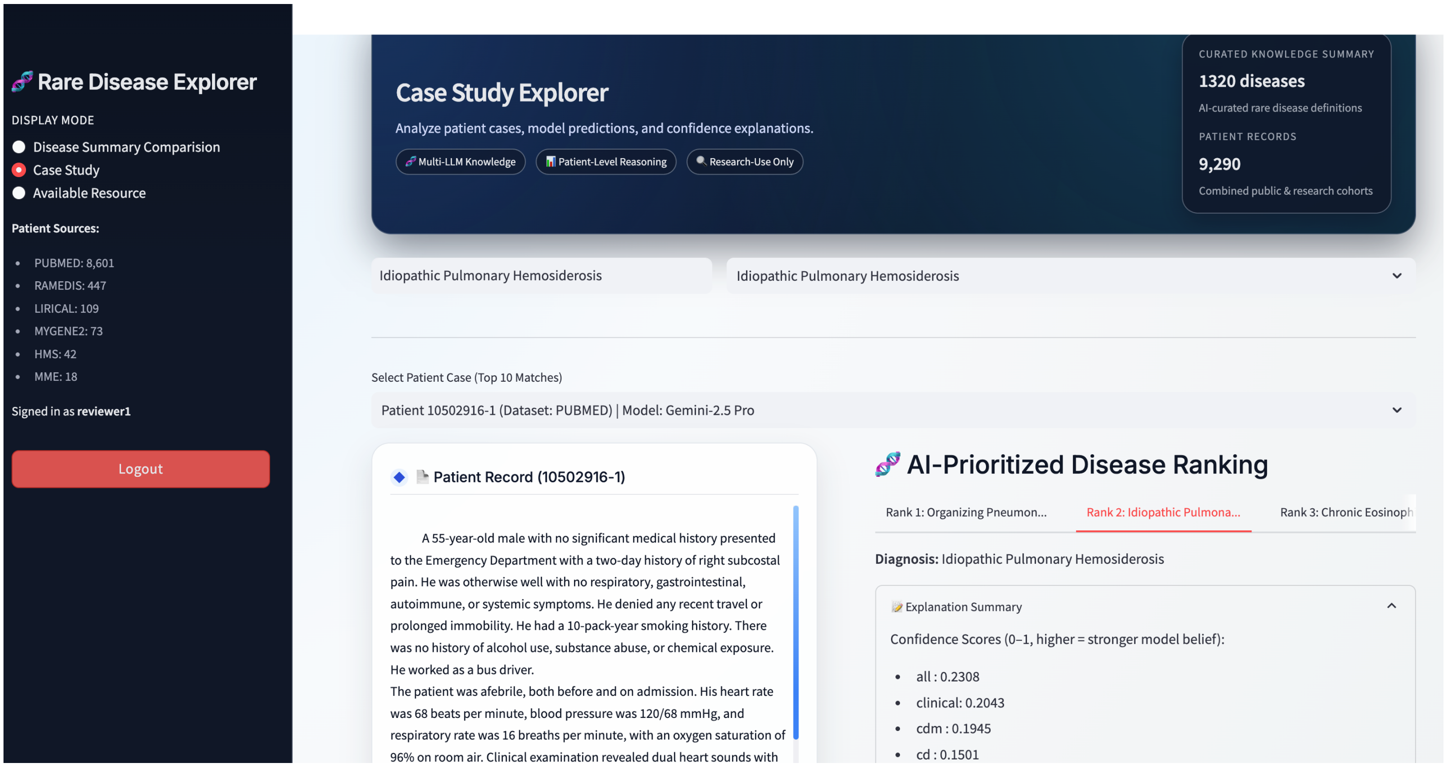


**Supplementary Fig. 2: Screenshot of one example within the public benchmarks, as well as the disease rankings produced by GEN-KnowRD.**

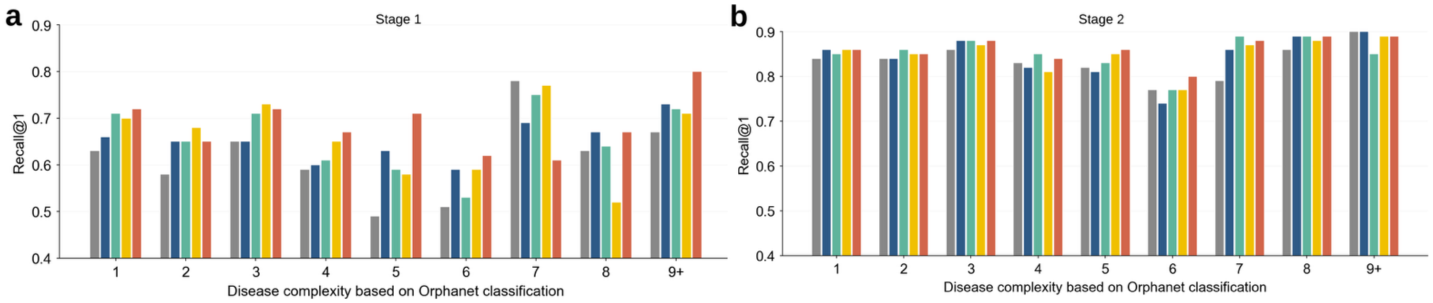


**Supplementary Fig. 3: Stage-wise evaluation of GEN-KnowRD's general-purpose rare disease screening across disease complexity. a,** Recall@1 of disease rankings out of Stage 1 across disease complexity categories based on Orphanet classification. **b,** Recall@1 of disease rankings out of Stage 2 across disease complexity categories based on Orphanet classification.

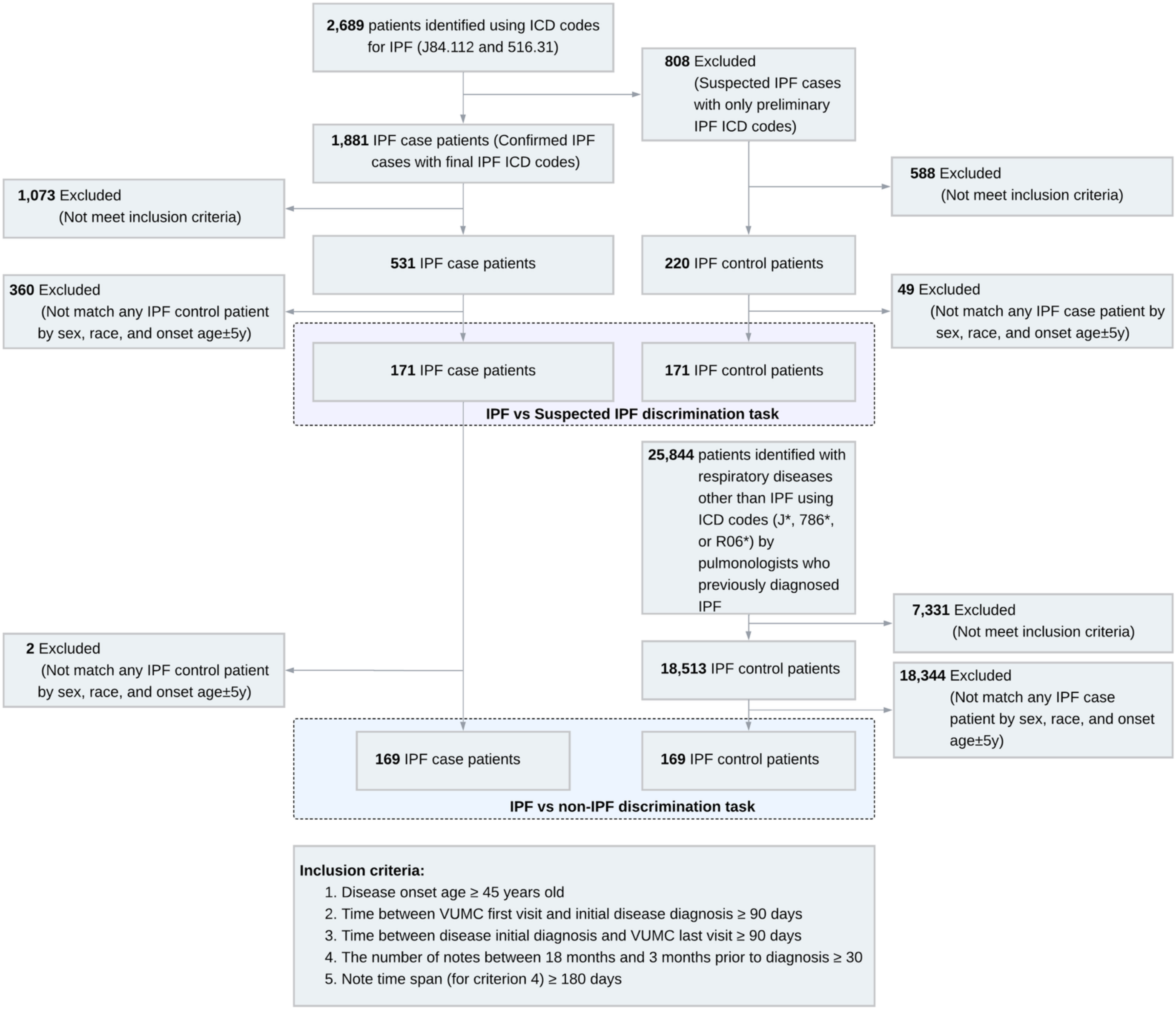


**Supplementary Fig. 4: Flow diagram illustrating patient inclusion and exclusion for the specialized rare disease discrimination tasks using longitudinal electronic health records at Vanderbilt University Medical Center.** Two tasks and the corresponding patient cohorts are constructed for idiopathic pulmonary fibrosis (IPF) early diagnosis: 1) IPF vs non-IPF and 2) IPF vs suspected IPF. Suspected IPF cases are defined as patients who were assigned with preliminary IPF ICD code and without final IPF ICD codes documented in the EHR.

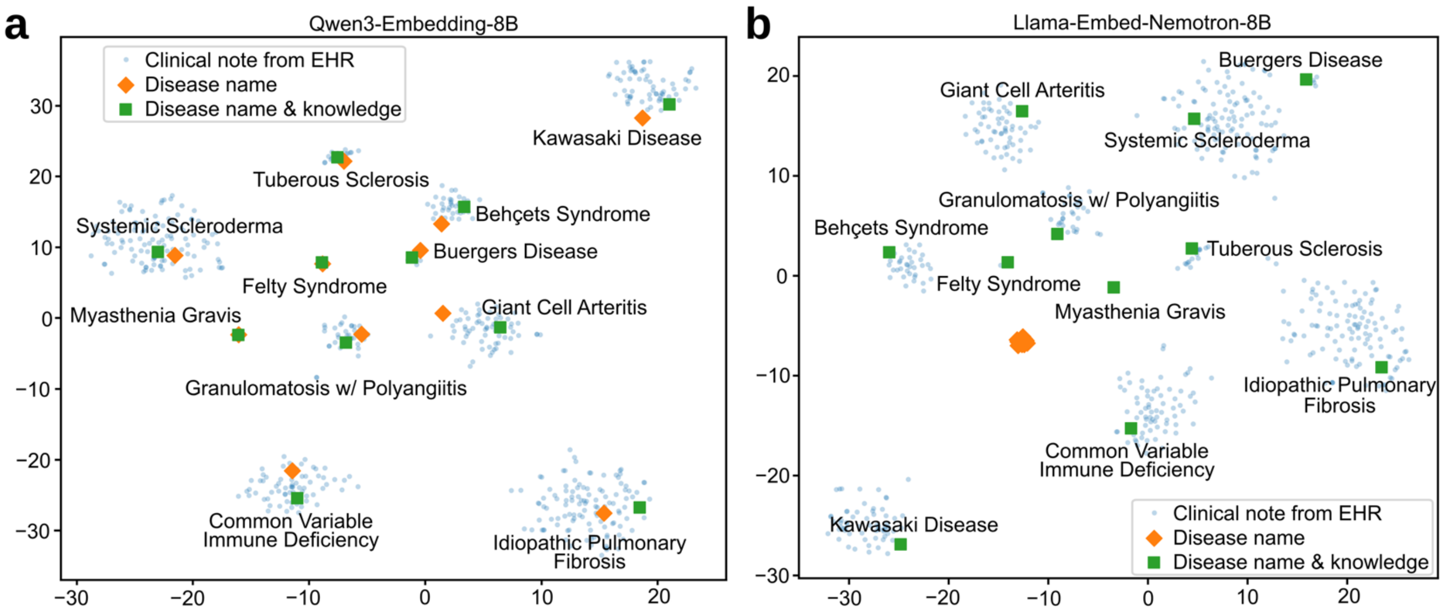


**Supplementary Fig. 5: T-SNE visualizations of diseases, represented by disease names and core sections from the Claude Sonnet 4-generated disease profiles, along with clinical notes from patients diagnosed with the corresponding diseases.** Embedding models used are **a,** Qwen3-Embedding-8B, and **b,** Llama-Embed-Nemotron-8B. Patient cohorts and the corresponding clinical notes for the 11 included diseases are constructed at Vanderbilt University Medical Center.

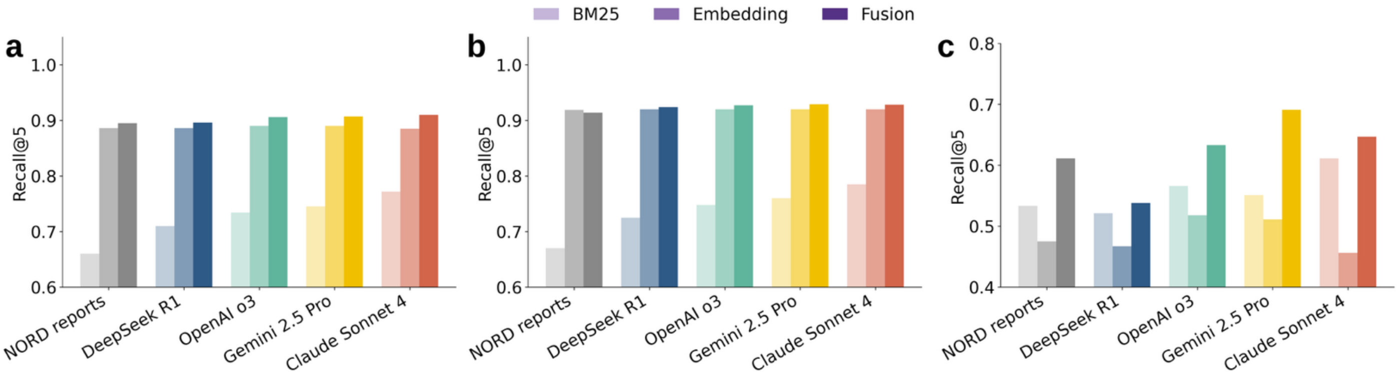


**Supplementary Fig. 6: Recall@5 for candidate disease rankings produced by BM25, embedding-based retrieval using Qwen3-Embedding-8B, and their fused ranking. a,** All benchmarks combined. **b,** PMC benchmark. **c,** Non-PMC benchmark.

**Supplementary Table 1: Prompt template for collecting rare disease profiles from large language models.**

You are an expert medical researcher. Your task is to generate a comprehensive and authoritative summary of _____, using the guidance provided within the triple quotation marks. This summary will serve as a reliable Wikipedia entry for use by medical researchers and healthcare professionals. In the "Clinical Presentation" section, you must include exhaustive information to guide clinical assessment. Ensure that all information is well-grounded in verified sources. Only use free-text descriptions with complete sentences. Do not use any other format. Again, you need to be as comprehensive as possible. Otherwise, you will be penalized.

"""

1 Disease Overview
- Definition: Provide a lay summary description of the disorder.
- Key Features: List hallmark clinical characteristics (≤ 3 phrases).
- Disease Category/Class: Identify broad classification (e.g., autoimmune, genetic, fibrotic lung disease).

2 Synonyms & Abbreviations
- Alternate Names: List all official and historical names.
- Preferred Acronym(s): Record standard abbreviations.

3 Subtypes / Variants
- Subdivisions: Enumerate recognized forms or phenotypes; note distinguishing criteria (age, severity, genetic marker, etc.).
- If none: State "No formal subtypes reported."

4 Epidemiology
- Prevalence / Incidence: Quantify (rate per population) and specify data source year/country.
- Demographics: Capture typical age of onset, sex ratio, ethnic or geographic clustering.
- Rarity Status: Indicate if classified as "rare" (≤ 200 K in U.S. or regional equivalent).

5 Etiology & Pathogenesis
- Primary Cause(s): Describe genetic mutation, autoimmune trigger, infection, toxin, or "idiopathic."
- Inheritance Pattern (if genetic): State autosomal dominant/recessive, X-linked, mitochondrial, etc.
- Pathophysiologic Mechanism: Summarize how the cause produces tissue damage or dysfunction.
- Key Risk Factors: List established environmental or lifestyle contributors.

6 Clinical Presentation
- Core Signs & Symptoms: Provide a comprehensive list, starting with most common/early and including late signs and symptoms.
- Progression Pattern: Detail typical progression patterns (acute, relapsing, chronic progressive).
- Variability between patients: Describe known variability in presentation.
- Major Complications: Note life-threatening or disabling sequelae.

7 Diagnostic Evaluation
- Clinical Criteria: Summarize key bedside findings required for diagnosis.
- Laboratory Tests: Specify biomarkers, antibody assays, enzyme levels, etc.
- Imaging / Instrumental Tests: List radiology, electrophysiology, biopsies essential for confirmation.
- Genetic Testing (if applicable): State recommended gene panels or specific variant analysis.
- Formal Guidelines: Reference any published diagnostic criteria sets.

8 Management & Standard Therapy
- First-Line Treatments: Name drugs, doses (range), or procedures routinely recommended.
- Second-Line / Adjunctive: List options for refractory or severe disease.
- Supportive Care: Include rehabilitation, nutritional guidance, devices, or monitoring protocols.
- Preventive Measures: Note prophylactic strategies (vaccines, lifestyle modifications).

9 Investigational / Emerging Therapies
- Therapies in Trials: Summarize novel agents, biologics, gene or cell therapies under clinical investigation.
- Trial Resources: Provide registry links or identifiers when available.

10 Prognosis
- Natural History: Describe typical survival or remission expectations without treatment.
- Impact of Therapy: State how modern treatment alters outcomes.
- Prognostic Factors: List variables associated with better or worse course.

"""

**Supplementary Table 2: Top-1 semantic matching rate for considered embedding models.**

| Model | Using disease name | Using disease name and core sections of disease profiles | | | |
|---|---|---|---|---|---|
| | | Claude Sonnet 4 | Gemini 2.5 Pro | OpenAI o3 | DeepSeek R1 |
| Llama-Embed-Nemotron-8B | 71.1% | - | - | - | - |
| Qwen3-Embedding-8B (Finetuned) | 84.7% | 87.1% (+2.4%) | 87.8% (+3.1%) | 86.1% (+1.4%) | 87% (+2.3%) |
| Qwen3-Embedding-8B | | 86.4% (+1.7%) | 84.6% (-0.1%) | 85.8%(+1.1%) | 82.7% (-2%) |

**Supplementary Table 3: Cohort characteristics for specialized IPF discrimination tasks.**

| | Task 1: IPF vs Non-IPF | | Task 2: IPF vs Suspected IPF | |
|---|---|---|---|---|
| | **Case** | **Control** | **Case** | **Control** |
| **# of patients** | 169 | 169 | 171 | 171 |
| **Gender** | 77 F; 92 M | 77 F; 92 M | 77 F; 94 M | 77 F; 94 M |
| **Race** | 11 B; 158 W | 11 B; 158 W | 11 B; 160 W | 11 B; 160 W |
| **Disease onset age (mean [STD])** | 70.37 [9.38] | 69.74 [9.03] | 70.30 [9.35] | 69.55 [9.39] |

*Pre-diagnosis window: time window between 18 and 3 months prior to the initial diagnosis.

**Supplementary Table 4: Prompt used to collect disease rankings from OpenAI GPT-5.**

```
You are an experienced specialist in diagnosing rare diseases.
You will be given a clinical case description. Your task is to generate a prioritized differential diagnosis consisting of the Top 5 most likely diseases.

Important rules:
1. Only select disease names from the provided list below. Do not use any diseases outside this list.
2. Provide a ranked list of the top 5 diseases, from most likely (1) to least likely (5).
3. Return the output strictly in the following JSON format (no extra text, explanation, or commentary):
{{
  "1": "disease_name_1",
  "2": "disease_name_2",
  "3": "disease_name_3",
  "4": "disease_name_4",
  "5": "disease_name_5"
}}

Disease list:
{disease_list}

Now analyze the following patient case description (enclosed in triple quotation marks):
\"\"\"{case_data}\"\"\"
```

**Supplementary Table 5: Comparisons of Recall@1 between GEN-KnowRD-Ensemble and GEN-KnowRD with individual knowledge sources.** *p* values are calculated using a two-sided Wilcoxon signed-rank test with Holm-Bonferroni correction.

| Dataset | GEN-KnowRD-Ensemble | GEN-KnowRD | | Adjusted *p* value |
|---|---|---|---|---|
| | Mean Recall@1 | Knowledge source | Mean Recall@1 | |
| Combined | 0.870 | NORD reports | 0.837 | <0.001 |
| | | DeepSeek R1 | 0.849 | <0.001 |
| | | OpenAI o3 | 0.858 | <0.001 |
| | | Gemini 2.5 Pro | 0.853 | <0.001 |
| | | Claude Sonnet 4 | 0.860 | <0.001 |
| PMC | 0.892 | NORD reports | 0.860 | <0.001 |
| | | DeepSeek R1 | 0.872 | <0.001 |
| | | OpenAI o3 | 0.880 | <0.001 |
| | | Gemini 2.5 Pro | 0.875 | <0.001 |
| | | Claude Sonnet 4 | 0.884 | <0.001 |
| Non-PMC | 0.586 | NORD reports | 0.546 | 0.028 |
| | | DeepSeek R1 | 0.553 | NS |
| | | OpenAI o3 | 0.575 | NS |
| | | Gemini 2.5 Pro | 0.578 | NS |
| | | Claude Sonnet 4 | 0.556 | 0.020 |

**Supplementary Table 6: UMLS concept categories used for concept extraction and the semantic types included within each category.**

| Concept Category | UMLS Semantic Types |
|---|---|
| Symptoms & Conditions | Disease or Syndrome<br>Sign or Symptom<br>Finding<br>Pathologic Function<br>Neoplastic Process<br>Mental or Behavioral Dysfunction<br>Congenital Abnormality<br>Anatomical Abnormality |
| Diagnostics & Laboratory Findings | Diagnostic Procedure<br>Laboratory Procedure<br>Laboratory or Test Result |
| Drugs & Procedures | Pharmacologic Substance<br>Clinical Drug<br>Therapeutic or Preventive Procedure<br>Organic Chemical |
| Genetics & Molecular Biology | Gene or Genome<br>Genetic Function<br>Amino Acid, Peptide, or Protein<br>Enzyme<br>Cell or Molecular Dysfunction |

**Supplementary Table 7: Expert evaluation criteria against disease profiles.**

**Overview**

This rubric is designed for clinical experts to systematically evaluate the quality of medical corpora describing rare diseases. Each dimension assesses a distinct aspect of corpus quality and should be scored holistically using the provided 5-point scale.

**Scoring Scale:**

- **1 = Unacceptable:** Fundamentally flawed; would cause harm if used
- **2 = Poor:** Significant deficiencies; requires major revision
- **3 = Adequate:** Meets minimum standards; some gaps present
- **4 = Good:** High quality with minor limitations
- **5 = Excellent:** Exemplary; meets the highest professional standards

**Evaluator Instructions**

- Read the entire corpus before scoring any dimension
- Score each dimension independently without reference to other dimensions
- Use the provided criteria strictly; do not interpolate between levels
- <u>Document specific reasons and examples justifying scores below 4</u>

**DIMENSION 1: FACTUAL ACCURACY**

**Definition:** The degree to which statements, data, and claims in the corpus are verifiable, correct, and consistent with current peer-reviewed evidence and established clinical guidelines.

**Scope of Evaluation:** This dimension evaluates the CORRECTNESS of information presented, not its completeness or practical usefulness. Consider whether pathophysiological mechanisms, epidemiological data, diagnostic criteria, therapeutic information, and medical terminology are accurate according to current scientific understanding and established consensus guidelines. Assess whether the corpus is free from factual errors, outdated information presented as current, or misleading statements.

| Score | Description |
|---|---|
| **5 - Excellent** | All information is accurate and reflects current evidence-based understanding. Terminology is precise and consistent. No factual errors detected. Appropriate qualification of uncertain or evolving knowledge. |
| **4 - Good** | Information is accurate with only minor imprecisions that would not affect clinical understanding or decisions. Terminology is largely correct with negligible inconsistencies. |
| **3 - Adequate** | Generally accurate but contains some errors or imprecisions that require correction. May include some outdated information or terminology inconsistencies, but core facts are sound. |
| **2 - Poor** | Contains notable factual errors or significantly outdated information that could mislead readers. Multiple terminology errors or inconsistencies present. |
| **1 - Unacceptable** | Contains fundamental factual errors, fabricated information, or dangerous misinformation that could lead to patient harm if used clinically. |

**Score: ___ / 5**

**Evaluator Notes:**

*Document specific factual errors or concerns identified:*

**DIMENSION 2: CLINICAL COMPLETENESS**

**Definition:** The extent to which the corpus covers all essential clinical domains necessary for comprehensive understanding and management of the disease, without critical omissions.

**Scope of Evaluation:** This dimension evaluates COVERAGE and COMPREHENSIVENESS, not accuracy or practical utility. Consider whether the corpus adequately addresses: disease definition and classification, etiology and risk factors, clinical presentation and natural history, diagnostic evaluation approaches, therapeutic management options, prognosis and disease course, complications and comorbidities, and considerations for special populations where relevant.

| Score | Description |
|---|---|
| **5 - Excellent** | Comprehensive coverage of all essential clinical domains. No significant omissions. Appropriate depth across all relevant aspects of the disease from definition through management and prognosis. |
| **4 - Good** | Covers all major clinical domains with only minor gaps in secondary areas. Essential information for understanding and managing the disease is present. |
| **3 - Adequate** | Covers most essential domains but has noticeable gaps in some areas. Core elements are present but certain important aspects receive insufficient attention. |
| **2 - Poor** | Significant gaps in coverage. Multiple essential clinical domains are missing or inadequately addressed, limiting the corpus's value as a comprehensive resource. |
| **1 - Unacceptable** | Critically incomplete. Major clinical domains are absent, rendering the corpus insufficient for meaningful understanding or management of the disease. |

**Score: ___ / 5**

**Evaluator Notes:**

*Identify critical content gaps:*

**DIMENSION 3: CLINICAL UTILITY**

**Definition:** The degree to which the corpus provides actionable, practically applicable information that supports clinical reasoning, decision-making, and patient care workflows.

**Scope of Evaluation:** This dimension evaluates USEFULNESS and APPLICABILITY for practicing clinicians, not accuracy or completeness alone. Consider whether the information enables clinicians to: recognize and diagnose the disease, make informed treatment decisions, stratify risk and prognosis, integrate knowledge into clinical workflow, communicate effectively with patients, and determine when specialist referral is needed.

| Score | Description |
|---|---|
| **5 - Excellent** | Highly actionable content that directly supports clinical decision-making. Information is presented in a manner that facilitates practical application. Enables confident diagnosis, treatment selection, and patient counseling. |
| **4 - Good** | Clinically useful with clear applicability to practice. Most information can be readily translated into clinical action with only minor gaps in actionable guidance. |
| **3 - Adequate** | Provides useful clinical information but requires additional interpretation or supplementation to guide specific decisions. Actionability is limited in some areas. |
| **2 - Poor** | Limited practical utility. Information is too theoretical, vague, or poorly organized to effectively support clinical decision-making without substantial external resources. |
| **1 - Unacceptable** | No meaningful clinical utility. Content cannot be practically applied to patient care or could lead to inappropriate clinical actions if followed. |

**Score: ___ / 5**

**Evaluator Notes:**

*Comment on practical applicability barriers:*

**DIMENSION 4: DISEASE SPECIFICITY**

**Definition:** The degree to which the corpus captures unique characteristics, distinguishing features, and context-specific considerations particular to the rare disease being described, avoiding generic or non-specific content.

**Scope of Evaluation:** This dimension evaluates SPECIFICITY and DIFFERENTIATION from similar or general conditions. Consider whether the corpus articulates pathognomonic or characteristic features, provides precise differential diagnosis guidance, describes disease-specific diagnostic and therapeutic approaches, addresses unique management considerations, acknowledges rare disease context challenges (diagnostic delays, limited evidence, orphan therapies, centers of excellence), and recognizes phenotypic heterogeneity within the condition.

| Score | Description |
|---|---|
| **5 - Excellent** | Content is highly specific to the target disease. Clearly articulates distinguishing features, disease-specific approaches, and unique considerations. Effectively differentiates from similar conditions. Appropriately addresses rare disease context. |
| **4 - Good** | Predominantly disease-specific with clear attention to distinguishing characteristics. Most content is tailored to the specific condition with only minor reliance on generic information. |
| **3 - Adequate** | Mix of disease-specific and generic content. Key distinguishing features are addressed but some sections could apply to related conditions without modification. |
| **2 - Poor** | Predominantly generic content with limited disease-specific information. Fails to adequately distinguish from similar conditions or address unique aspects of the target disease. |
| **1 - Unacceptable** | Content is largely generic or could apply to any similar condition. No meaningful disease-specific information. May conflate the target disease with related but distinct entities. |

**Score: ___ / 5**

**Evaluator Notes:**

*Comment on generic vs. disease-specific content balance:*

**GLOBAL ASSESSMENT**

**Overall Quality Rating (select one):**

| Rating | Description |
|---|---|
| ☐ **Unacceptable** | Contains errors or omissions that could result in patient harm; not suitable for any clinical or educational use. |
| ☐ **Below Standard** | Significant deficiencies requiring substantial revision before use; may be misleading to non-experts. |
| ☐ **Acceptable** | Meets minimum professional standards; suitable for general reference with appropriate caveats. |
| ☐ **Good** | High-quality content appropriate for clinical education and decision support with minor limitations. |
| ☐ **Excellent** | Exemplary content suitable for guideline development, specialist education, or authoritative reference. |

**Supplementary Table 8: Prompt used to re-structure the NORD rare disease reports for fair comparison.**

You are a rare disease expert.

Task:

Read the ORIGINAL article text about {disease} provided below, and ONLY move sentences from the original text into the 4 predefined categories. Use the full category definitions and requirements provided in the triple quotation marks.

Hard rules (must follow exactly):

1) Do NOT rewrite, paraphrase, shorten, merge, or expand any sentence. Copy sentences EXACTLY as they appear.
2) One sentence can appear in ONLY ONE category (choose the best single category).
3) If a sentence does not clearly fit any category, IGNORE it (do not include it anywhere).
4) Do NOT add any new sentences. The ONLY exception is:
   - If the original text contains no subtype/variant statements, then output exactly this single sentence under "Subtypes / Variants": "No formal subtypes reported."
   - Otherwise, do not output that sentence.
5) Preserve the original sentence punctuation, capitalization, and wording.
6) Output must be VALID JSON only (no markdown, no commentary, no extra keys).

Category definitions (apply these exactly):

"""
1. Subtypes / Variants

• Subdivisions: Enumerate recognized forms or phenotypes; note distinguishing criteria (age, severity, genetic marker, etc.).
• If none: State “No formal subtypes reported.”

2. Clinical Presentation

• Core Signs & Symptoms: Provide a comprehensive list, starting with most common/early and including late signs and symptoms.
• Progression Pattern: Detail typical progression patterns (acute, relapsing, chronic progressive).
• Variability between patients: Describe known variability in presentation.
• Major Complications: Note life-threatening or disabling sequelae.

3. Diagnostic Evaluation

• Clinical Criteria: Summarize key bedside findings required for diagnosis.
• Laboratory Tests: Specify biomarkers, antibody assays, enzyme levels, etc.
• Imaging / Instrumental Tests: List radiology, electrophysiology, biopsies essential for confirmation.
• Genetic Testing (if applicable): State recommended gene panels or specific variant analysis.
• Formal Guidelines: Reference any published diagnostic criteria sets.

4. Management & Standard Therapy

• First-Line Treatments: Name drugs, doses (range), or procedures routinely recommended.
• Second-Line / Adjunctive: List options for refractory or severe disease.
• Supportive Care: Include rehabilitation, nutritional guidance, devices, or monitoring protocols.
• Preventive Measures: Note prophylactic strategies (vaccines, lifestyle modifications).
"""

JSON output schema (must follow exactly):

- Keys must be exactly:
  "Subtypes / Variants"
  "Clinical Presentation"
  "Diagnostic Evaluation"
  "Management & Standard Therapy"

- Each value must be an array of strings.
- Each string must be one verbatim sentence copied from the original text.
- No additional keys.

Original article text:

"""{article_text}"""

Now perform the task for {disease}. Return JSON only.
